# Assessing the effect of social contact structure on the impact of pneumococcal conjugate vaccines

**DOI:** 10.1101/2024.08.13.24311931

**Authors:** Anabelle Wong, Sarah C. Kramer, Daniel M. Weinberger, Matthieu Domenech de Cellès

## Abstract

Although pneumococcal conjugate vaccines (PCVs) have greatly reduced diseases caused by vaccine-targeted serotypes (VT) of *Streptococcus pneumoniae*, vaccine impact may be eroded by the increase in rates of disease caused by non-vaccine serotypes (NVT). Here, we investigated the effect of social contact patterns on the dynamics of vaccine impact in carriage.

We developed a neutral, age-structured, Susceptible–Colonized model incorporating VT-NVT co-colonization and verified it against real-world carriage data in children. Using contact matrices from 34 countries, we assessed the impact of contact patterns on the time required to eliminate VT (i.e., 95% reduction in VT proportion in carriage). Finally, we quantified the contribution of various parameters—such as vaccine efficacy, coverage, immunity waning, and population susceptibility—to the dynamics of VT elimination.

Our model recapitulated the observed decline of VT carriage and showed that varying the contact structure alone led to different time-to-elimination. We found that higher total contact rate and assortativity in children under 5 accelerated VT elimination. Additionally, higher vaccine efficacy and coverage, and slower immunity waning led to shorter time-to-elimination.

These findings illuminate the mechanisms controlling the dynamics of vaccine impact and may help predict the impact of PCVs in communities with different contact patterns.

## Introduction

*Streptococcus pneumoniae* is one of the five leading pathogens for the estimated 7.7 million bacteria-associated deaths globally [1]. The first several generations of pneumococcal conjugate vaccines (PCVs) have reduced invasive pneumococcal disease (IPD) substantially in all age groups [2]. However, the reduction in rates of disease caused by vaccine-targeted serotypes of pneumococci (VT) was partially offset by an increase in rates of disease caused by non-vaccine-targeted serotypes (NVT) [3,4]. This phenomenon, known as “serotype replacement”, occurred because PCVs targeted a subset of over 100 identified serotypes [5,6], reducing the fitness of VT and changing the competitive balance between VT and NVT [7,8]. Nasopharyngeal carriage is a prerequisite for pneumococcal diseases, and the reduction in carriage in immunized children leads to indirect protection of unvaccinated children and adults [9]. Likewise, serotype replacement in carriage may erode the population-level impact of PCV and thus demands public health attention.

Observed serotype replacement in diseases was initially more pronounced in the UK than in the US, for which multiple possible explanations have been suggested: the distribution of risk factors, the vaccination schedule and coverage, and the pre-PCV composition of circulating serotypes [10]. While replacement in diseases is partial, replacement in carriage is almost complete, and it occurs faster in some populations than others [3,11,12]. However, the mechanisms driving such variation remain unclear. One potential determinant for the serotype replacement dynamics is the social contact structure in a population. Carriage studies have shown that social contact with preschool-age children is associated with higher prevalence of pneumococcal carriage [13,14]. While social contact structures are thought to be major drivers of infectious disease dynamics [15], there have not been studies investigating the effect of social contact structure on the dynamics of vaccine impact in pneumococcal carriage. Addressing this knowledge gap can elucidate the potential mechanisms controlling the dynamics of vaccine impact and serotype replacement and may help predict the impact of the higher-valency PCVs in communities with different contact patterns.

In this study, we developed a mathematical model parameterized with empirical data to simulate the dynamics of serotype changes after PCV introduction (Figure 1) and verified it against observed prevalence of VT carriers among children in France, Alaska (US), Massachusetts (US), and the UK. Then, using contact matrices from 34 countries empirically inferred by [16], we interrogated the impact of social contact patterns on the trajectory of VT carriage decline (Figure 2). In addition, we quantified the effect of key parameters such as vaccine efficacy and population susceptibility by changing one parameter at a time. Our findings showed that variations in social contact structure alone led to different time-to-elimination. We found high association between the contact pattern features in children under 5 and time-to-elimination. More broadly, our findings highlight the need to consider social contact structure when assessing the impact of vaccines.

**Figure 1.**
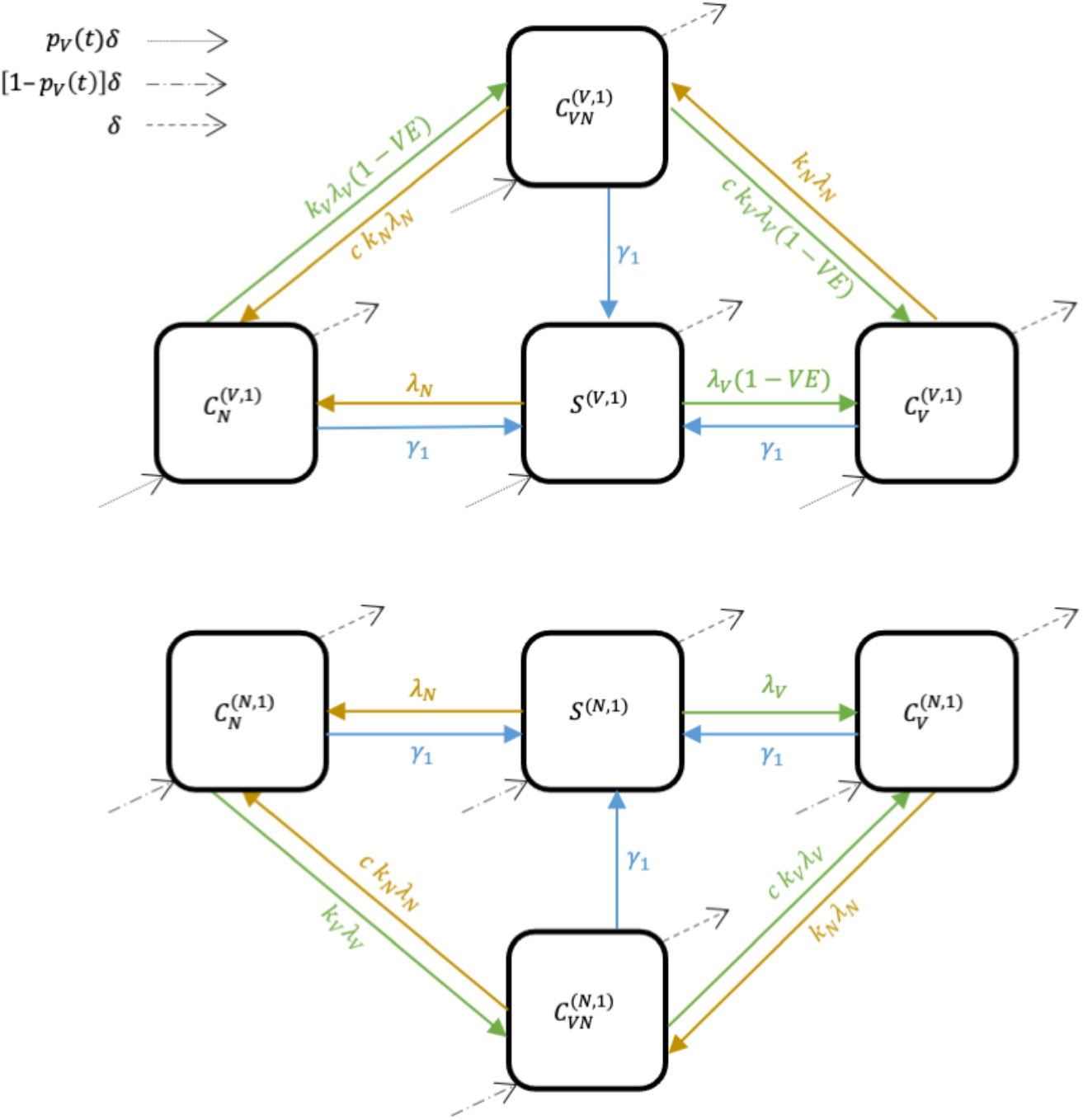
A neutral, age-structured, Susceptible–Colonized transmission model. Boxes represent the state variables (*S*–Susceptible, *C*–Colonized; superscripts indicate vaccine status and age: *V* –vaccinated, *N* –unvaccinated; subscripts indicate the colonizing serotype: *V* – vaccine-targeted serotypes (VT), *N*–non-vaccine serotypes (NVT), *VN*–both VT and NVT). Arrows represent the movement of individuals between states (solid arrows: green–due to colonization with VT, brown–due to colonization with NVT, blue–due to clearance of colonizing serotypes; dotted arrow: due to aging from age 0 and being vaccinated; dot-dash arrow: due to aging from age 0 and not being vaccinated, dashed arrows: due to aging). For simplicity, only the second age group is represented.

**Figure 2.**
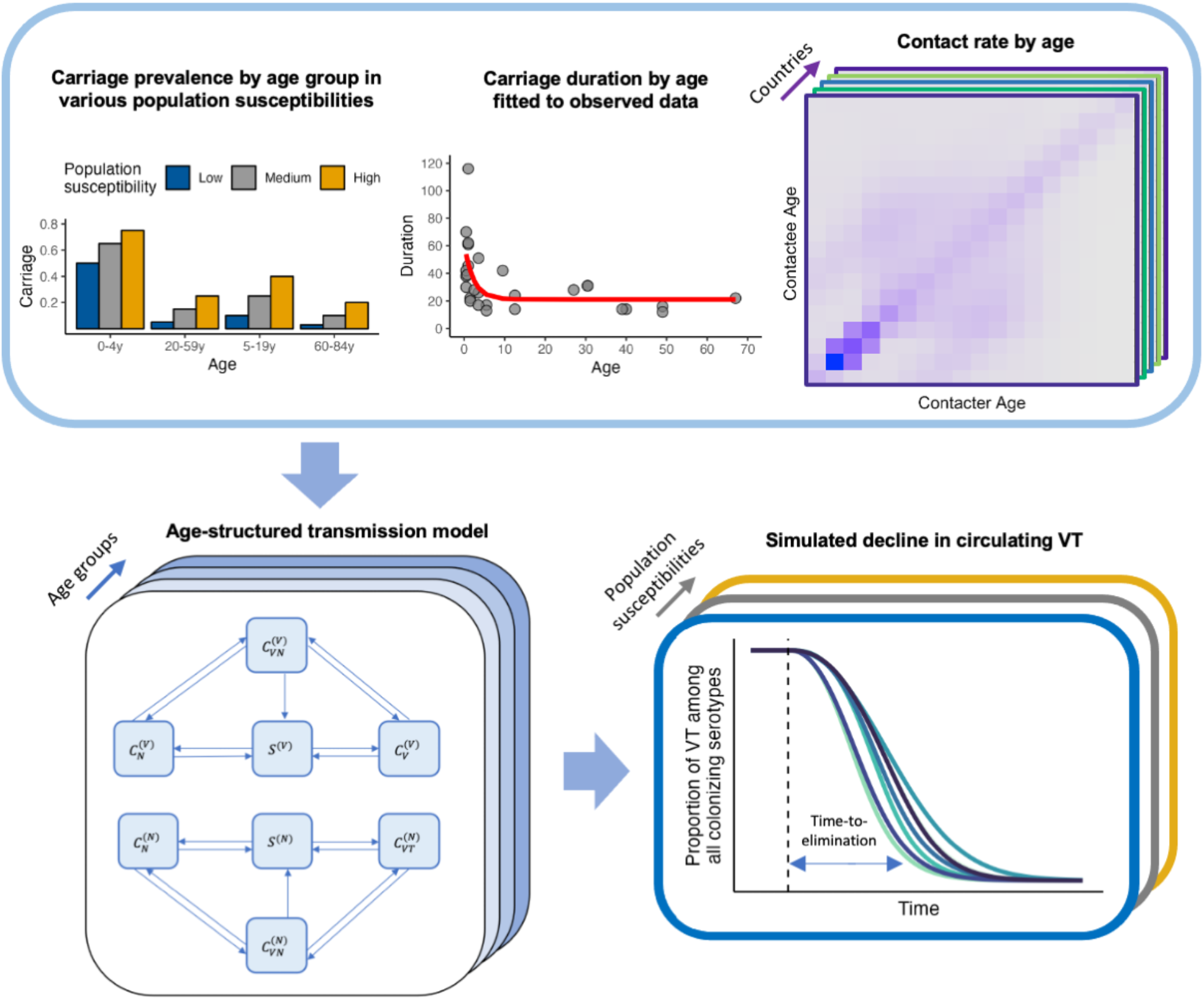
The modeling workflow. The top row shows the components entering the transmission model, from left to right: overall carriage prevalence by age group under different population susceptibilities, carriage duration by age fitted (red line) to observed data (grey points), and contact matrices from various countries. The bottom left panel shows the age-structured, Susceptible–Colonized transmission model. The bottom right panel shows the simulated decline in the proportion of VT among circulating serotypes; blue double arrow indicates the outcome – time-to-elimination – defined as the time between vaccine introduction (dashed line) and the time point when the proportion of VT among circulating serotypes dropped to 5% of its initial value in age 0.

## Results

### Real-world parameter sets allow the model to reproduce observed VT-carrier prevalence in children

We formulated a deterministic, Susceptible–Colonized model that simulates the transmission of VT and NVT carriage before and after the introduction of PCVs. The model was an instance of neutral null models proposed by Lipsitch et al. for multistrain pathogens [17]. A key property of these models is the lack of a stable coexistence equilibrium, so that any initial level of coexistence will be maintained over time for identical strains. While individual pneumococcal serotypes differ in fitness [18], there is no conclusive evidence for differential transmissibility or duration of carriage for VT and NVT [19]. In addition, pneumococcal diversity and fitness differences extend beyond the serotype level [12]. Given these considerations, we opted for a neutral model as a parsimonious way to achieve initial levels of co-existence without having to specify serotype-specific parameters.

The simulations using location-specific parameter sets (Table 1) broadly captured the observed dynamics of VT-carrier prevalence in children in the post-PCV era in the UK, Alaska (US), and Massachusetts (US), and with some discrepancy, in France (Figure 3). In general, the observed VT-carrier prevalence declined slightly more rapidly than in the simulation. The VT-carrier prevalence in the pre-vaccine era was higher in France (43.9%, 95% CI 38.4–49.4%) [20] and the UK (31.9%, 28.1–36.1%) [21] than in Alaska (20%, 15.7–24.7%) [22]. For Massachusetts, the VT-carrier prevalence was 9.7% half year after vaccine introduction [23]. In all locations, the rapid decline in VT-carriers immediately after vaccine introduction was followed by a slower decline as VT-carriers became less prevalent.

**Table 1.** Observed carriage and parameter set from four locations.

| **Location** | **Sample characteristics** | **Overall carriage (age 0, 1-4, 5-17, 18-39, 40-59, 60-84)** | **Initial proportions of VT-, NVT-carriers** | **Vaccine coverage** |
| --- | --- | --- | --- | --- |
| France [20] | Children 3–40 months attending daycare center | 0.59, 0.59, 0.30, 0.10, 0.10, 0.10 [20,26] | 0.75, 0.25 [20] | 2004-05: 61%<br>2005-05: 74%<br>2006-07: 86%<br>2007-08: 90% [33] |
| UK [21] | Children 1–5 years attending primary care practices | 0.49, 0.49, 0.21, 0.08, 0.08, 0.08 [21] | 0.659, 0.341 [21] | 90% [58] |
| Alaska, US [22] | Children 3 months–5 years attending primary care practices | 0.38, 0.38, 0.30, 0.10, 0.10, 0.10 [22,26] | 0.53, 0.47 [22] | 60% [22] |
| Massachusetts, US [23] | Children 3 months–7 years attending primary care practices | 0.28, 0.28, 0.28, 0.10, 0.10, 0.10 [23,26] | 0.36, 0.64 [23] | 85% [23] |

**Figure 3.**
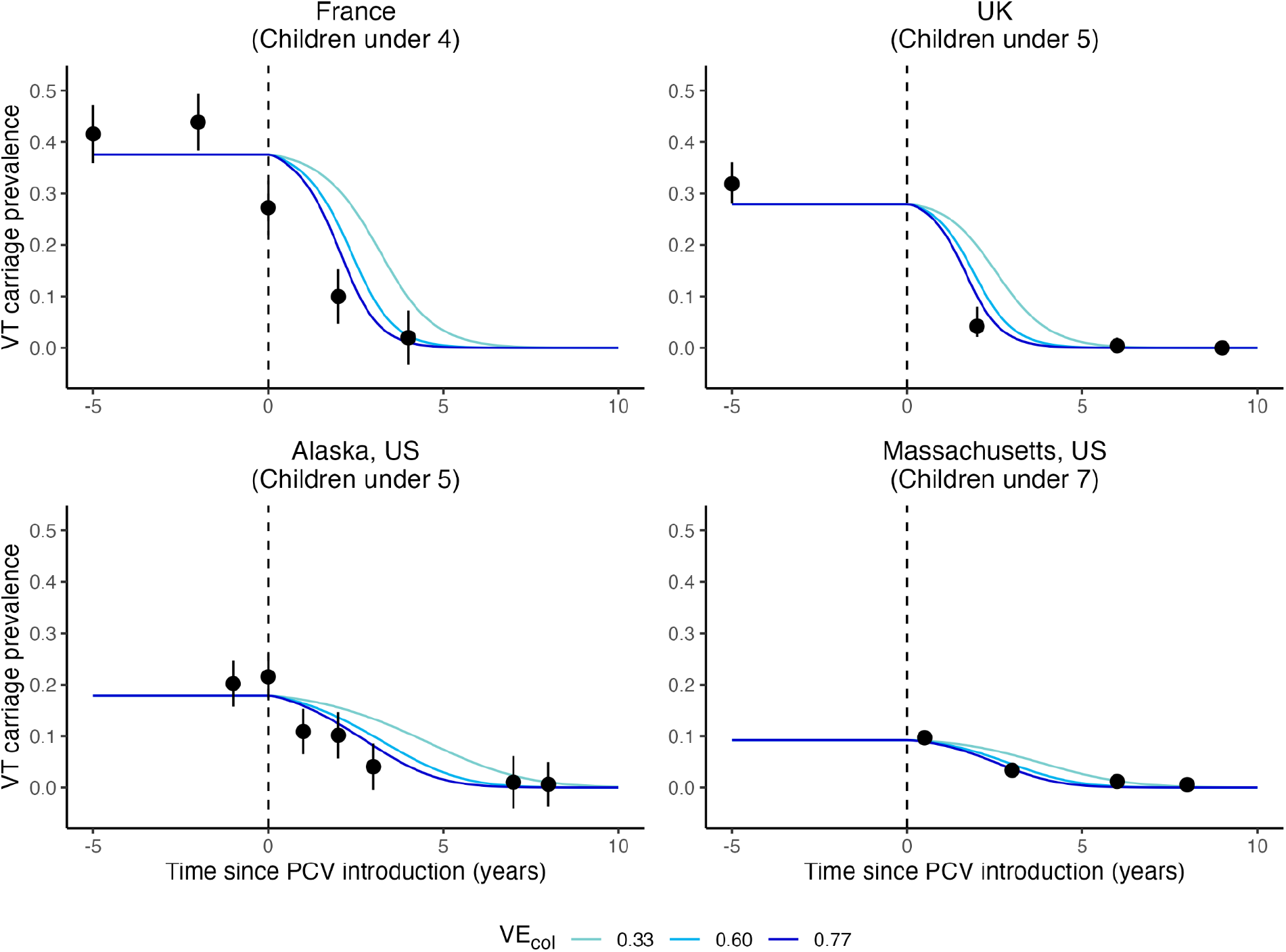
Simulated VT-carrier prevalence in children versus observed data in four locations. The lines indicate the simulated VT-carrier prevalence in children using a range of assumed vaccine efficacies against colonization acquisition (VE_col_) (light blue: 0.33, blue: 0.60, dark blue: 0.77) in four locations: France (top left), UK (top right), Alaska, US (bottom left), and Massachusetts, US (bottom right) from before to after the introduction of the pneumococcal conjugate vaccines (dashed line). Black points show the observed VT-carrier prevalence with 95% CI indicated by the error bars.

### The time-to-elimination was predicted to be shortest in children aged 1–5

We defined time-to-elimination as the duration between vaccine introduction and the time point when replacement was considered complete (i.e., 95% reduction in VT proportion in carriage). Using contact matrices derived from census and survey data in 34 countries [16], our transmission model produced variable time-to-elimination, ranging from 3.8 to 6 years in newborns, which was a fully unvaccinated age population and thus reflected the indirect effect of PCV introduction. The time-to-elimination in adults was similar to that in age 0 (Figure 4). In contrast, the time-to-elimination was the shortest in children of age 1 and above until age 5 in most countries and until age 10–11 in Ireland, the Netherlands, and the US. This finding corresponded well with the observation that PCV impact could be observed earlier in children than in adults [24], which is likely due to children of these ages having received the vaccine themselves and benefiting from both direct and indirect protections. They were also the age populations with the highest VT-carrier prevalence (Supplementary Figure 4) and contact rate (Figure 5A).

**Figure 4.**
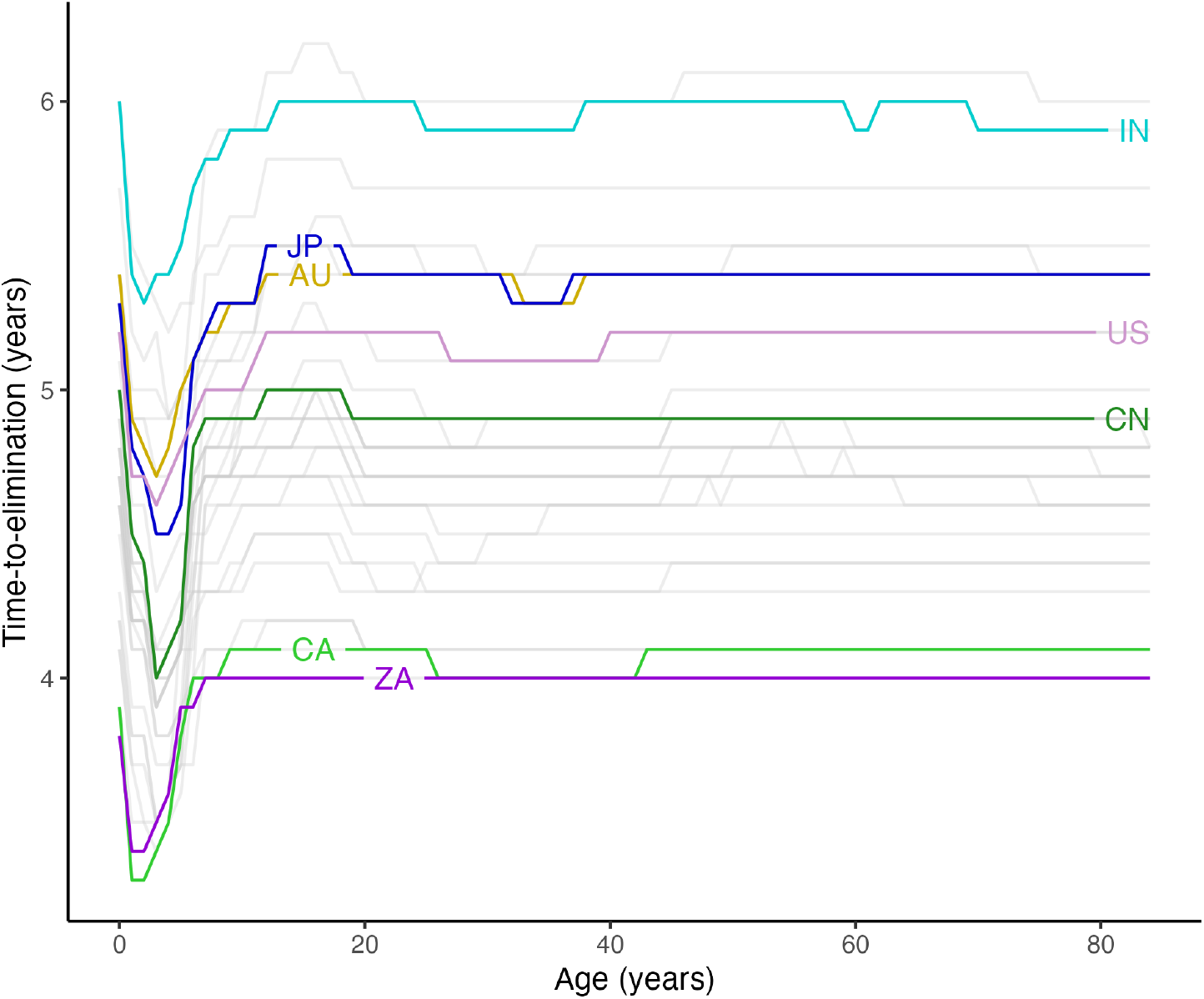
The predicted time-to-elimination by age in 34 countries. The lines show the simulated time-to-elimination by age using the contact matrices from 34 countries. The results for Australia (AU, yellow), Canada (CA, light green), China (CN, green), India (IN, light blue), Japan (JP, blue), South Africa (purple), and the United States (light purple) are highlighted as examples.

**Figure 5.**
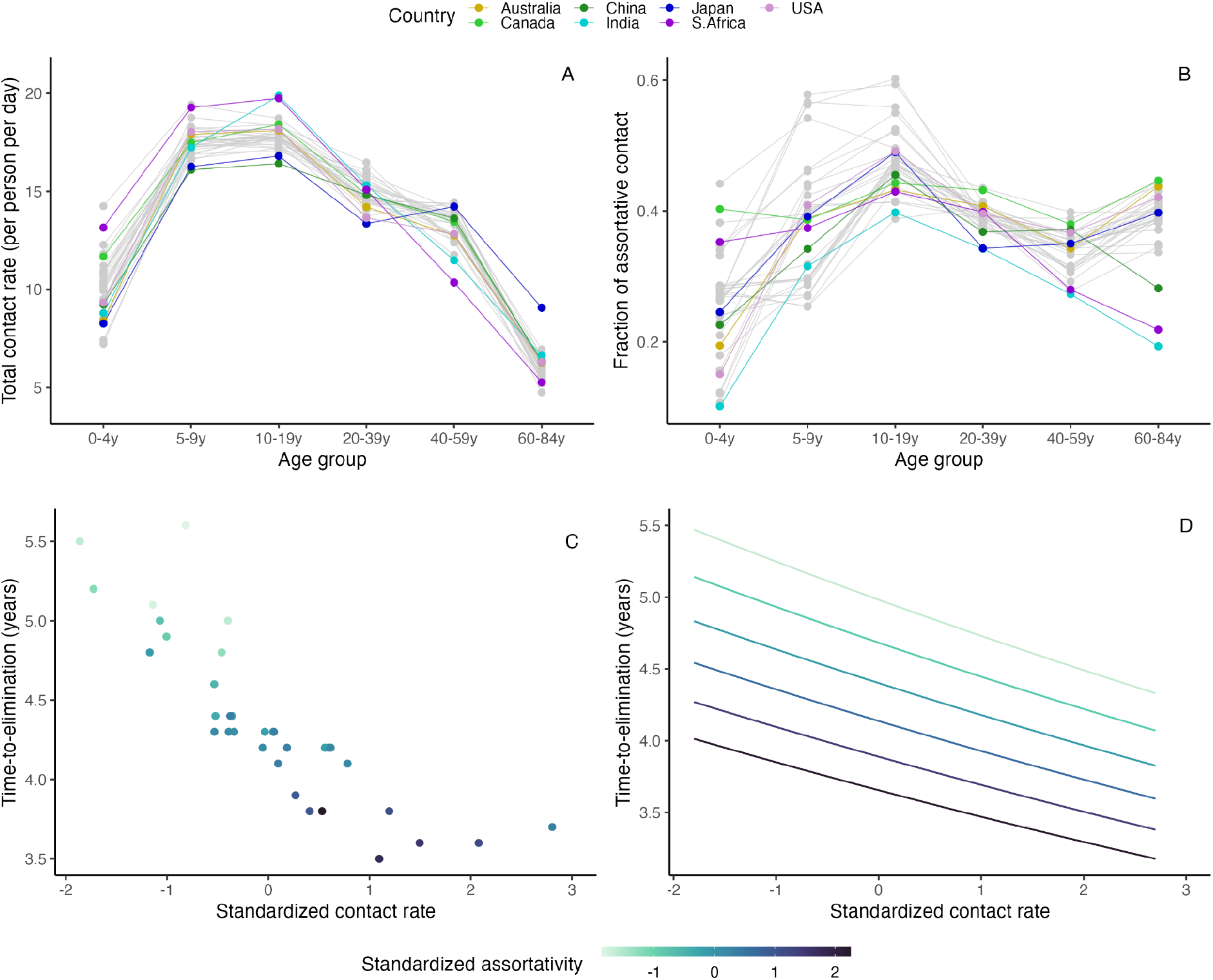
Total contact rate and assortativity predict time-to-elimination. The top row shows two contact features by age group in 34 countries: total contact rate, defined as the average total daily contacts in the age group (A), and assortativity, defined as the fraction of within-age group contact (B). The data points from Australia (yellow), Canada (light green), China (green), India (light blue), Japan (blue), South Africa (purple), and the United States (light purple) are highlighted as examples. The bottom row shows the correlation between time-to-elimination and standardized contact rate (x-axis) by standardized assortativity (color scale) in children under 5 in the simulated data (C) and the generalized linear model (D).

In the sensitivity analyses, we considered two scenarios: (1) a lower prevalence of carriers at age 0 due to the time lag from birth to first pneumococcal acquisition, and (2) a higher prevalence of carriers in all ages to simulate settings with higher pneumococcal burden (Supplementary Figure 4). The results remained similar (time-to-elimination range: 4.2–7.1 years, 4.4–6.9 years).

### Time-to-elimination was highly dependent on contact patterns in children under 5

To delineate the effect of mixing patterns in different age groups, we looked at two age group-specific social contact features that may be important for respiratory infection transmission [25]: contact rate (total daily contacts) and assortativity (fraction of within-group contact).

Across countries, the contact rate increased from children under 5 (0–4y) to peak around school-age (5–9y) and teenage years (10–19y), and declined towards older age (65–84y) (Figure 5A). In general, assortativity tended to be the lowest in children under 5. The change of assortativity with age was more variable than that of contact rate across countries, and we noted four major patterns (Figure 5B). In some contact matrices (e.g., Australia, China, India, Japan, USA), the fraction of assortative contact increased with age, reaching the peak among teenagers, and either remained high (e.g., Australia, Japan, USA) or declined (e.g., China, India) in adulthood. In other contact matrices (e.g., Canada, South Africa), assortativity were similar from birth until teenage and then either remained (e.g., Canada) or declined towards older age (e.g., South Africa).

In our simulation, we assumed children under 5 had the highest prevalence of carriers based on a systematic review [26]. However, this age group bore lower contact rates than the other age groups (Figure 5A). In contrast, age groups with higher contact rates (5–39y) tended to have lower carriage prevalence (Figure 5A, Supplementary Figure 4).

A plot of simulated time-to-elimination against contact rate and assortativity revealed a strong negative correlation between the contact patterns and time-to-elimination in children under 5 but not in other age groups (Figure 5C, Supplementary Figure 9). Since time-to-elimination in all ages was correlated within the same country (Figure 4), this result suggests the contact rate and assortativity in children under 5 may be useful in predicting the time-to-elimination in a country. Using a generalized linear model (GLM) with only these two predictors, we found that contact rate and assortativity in children under 5 explained most of the variability in the simulated time-to-elimination (R-squared of 0.95). Both features accelerated reduction of VT (Figure 5D): one standard deviation of increase in total contact rate and fraction of assortative contact shortened time-to-elimination by 5.2% (95%CI 3.7–6.7%) and 7.7% (95%CI 6.3–9.2%) respectively. To test the prediction performance of this model, we left 4 randomly selected contact matrices out as the test set and used the remaining 30 contact matrices as the training set. Repeating this procedure 10 times gave a mean relative absolute error (MRAE) of 1.2–5% (Supplementary Table 2), indicating good out-of-sample prediction. Hence, these two features of social contacts in children under 5 are useful predictors of time-to-elimination, highlighting the key role of this age group in the transmission dynamics of pneumococcal carriage.

**Table 2.** A list of parameters and their values in the model.

| Parameter | Interpretation | Value | Range tested | Source |
| --- | --- | --- | --- | --- |
| $\beta_V^{(i)} (= \beta_N^{(i)})$ | Age-specific susceptibility to carriage acquisition | $\beta^{(0,\dots,4)} = 0.015$<br>$\beta^{(5,\dots,19)} = 0.004$<br>$\beta^{(20,\dots,59)} = 0.003$<br>$\beta^{(60,\dots,84)} = 0.005$ | $\pm 20\%$ for high and low population susceptibility respectively | [59] |
| $i1/\gamma_i$ | Age-specific average duration of carriage | See Supplementary Figure 1 | / | Fitted to observed data (Supplementary Data 1) |
| $\delta_i$ | Aging rate | $1\text{yr}^{-1}$ | / | Each age group is 1 year |
| $k_N (= k_V)$ | Competition parameter: Effect of existing VT (NVT) carriage on acquiring NVT (VT) carriage | 0.5 | 0.1, 0.25, 0.75 | [18,45,46] |
| $c$ | Fraction of co-carriers returning to $C_V$ ($C_N$) upon reinfection with VT (NVT) | 0.5 | / | [17] |
| $q$ | Relative infectiousness with each serotype for co-carriers | 0.5 | / | [17] |
| $\epsilon_V$ | Vaccine efficacy against carriage acquisition | 60% | 33%, 77% | [50] |
| $p_V$ | Vaccine coverage | 90% | 50% | [11] |
| $\alpha_V$ | Waning rate of vaccine-conferred immunity | 0 per year | 0.1, 0.2, 0.3 per year | [52,53] |
| $f_C^{(i)}(0)$ | Initial prevalence of carriers in age group $i$ | $f_C^{(0,\dots,4)}(0) = 0.5$<br>$f_C^{(5,\dots,19)}(0) = 0.2$<br>$f_C^{(20,\dots,59)}(0) = 0.1$<br>$f_C^{(60,\dots,84)}(0) = 0.1$ | (i) Lower carriage prevalence in age 0,<br>(ii) Higher carriage prevalence in all ages<br>See Supplementary Figure 4 | [26],<br>observed data (Supplementary Data 2) |
| $f_V(0), f_N(0)$ | Initial proportions of VT-, NVT-carriers | $f_V(0): 0.6$<br>$f_N(0): 0.4$ | $f_V(0): 0.2-0.8$<br>$f_N(0): 0.2$ | Observed data (Table 1, Supplementary Data 3) |

### Higher vaccine efficacy and coverage and slower waning of vaccine immunity accelerate time-to-elimination

To investigate the effect of key parameters on time-to-elimination, we varied one parameter at a time and measured the simulated time-to-elimination using a subset of 7 contact matrices representative of all the contact matrices from [16]. The key parameters tested were vaccine efficacy against carriage acquisition, vaccine coverage in the target age group (1-year-old), waning rate of vaccine immunity, the initial proportion of VT-and NVT-carriers, and population susceptibility to carriage acquisition (with 3 levels considered: high, medium, or low). Table 2 shows the list of parameters in the model.

Among the key parameters studied, vaccine factors resulted in the most prominent changes in time-to-elimination. When vaccine coverage reached 90%, using a highly efficacious vaccine (vaccine efficacy: 77%) led to a 1.9–2.6-year reduction in time-to-elimination compared with a less efficacious vaccine (vaccine efficacy: 33%) (Figure 6A). At lower coverage (50%), VT elimination was slower (5.1–7.8 years vs. 3.8–6 years in 90% coverage), and the same increase in vaccine efficacy caused a greater reduction in time-to-elimination (Supplementary Figure 5). In addition to no waning, we tested various durations of vaccine-conferred immunity and found that rapid waning (immunity duration of 3 years) slowed elimination by 0.5–1.6 years compared with slow waning (immunity duration of 10 years) (Figure 5B).

**Figure 6.**
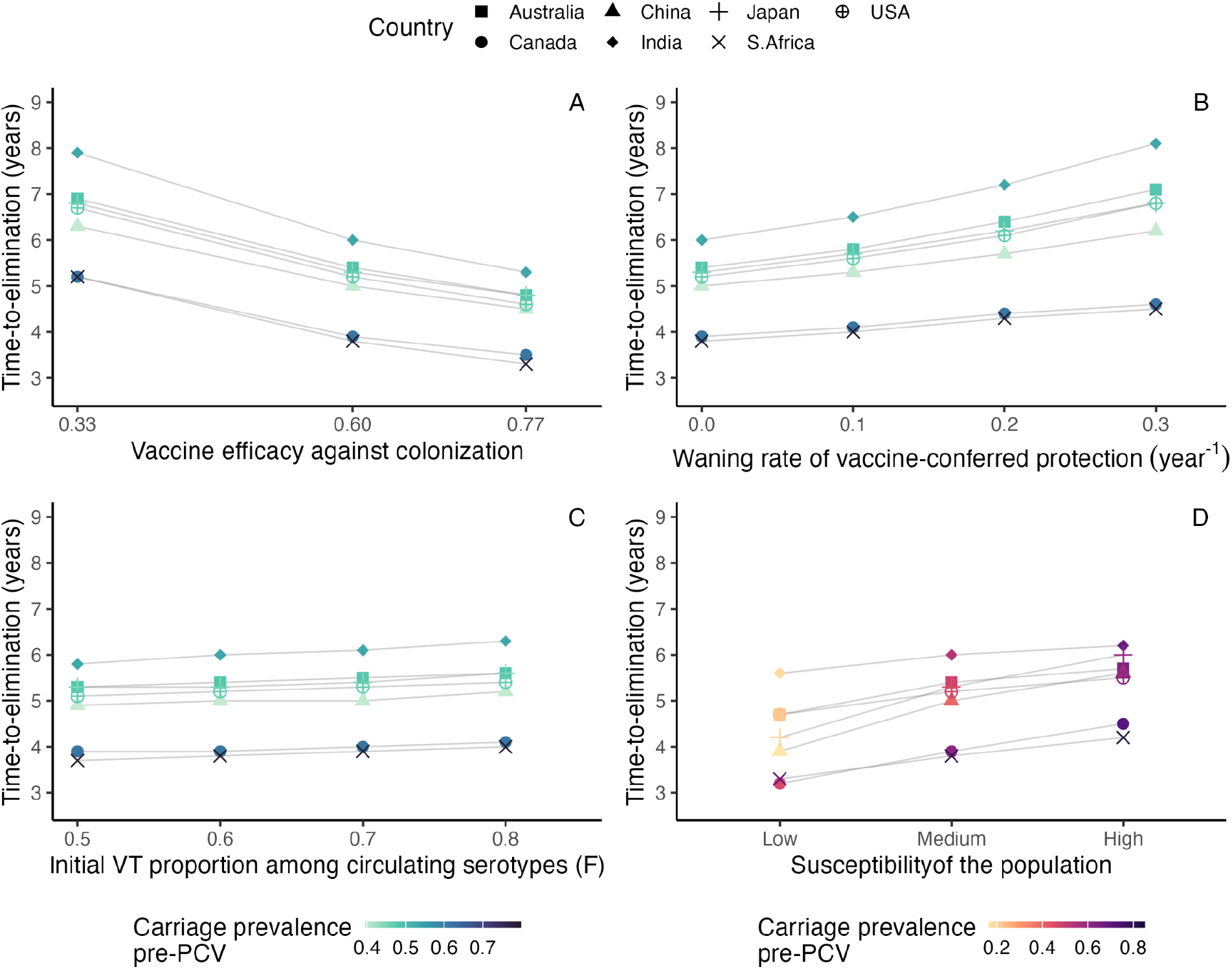
The effect of key parameters on time-to-elimination. Each point represents the time-to-elimination simulated by changing one key parameter at a time (A: vaccine efficacy against colonization acquisition, B: waning rate of vaccine-conferred immunity against colonization acquisition, C: initial proportion of VT among circulating serotypes, D: population susceptibility) using the contact matrices from 7 countries (square: Australia, circle: Canada, triangle: China, diamond: India, plus: Japan, cross: South Africa, plus in circle: the United States). The overall carriage prevalence before the introduction of the pneumococcal conjugate vaccines (PCV) in each country is represented by the same color scale in panels A, B, and C, and with a different color scale in panel D, because changing the population susceptibility naturally resulted in different pre-PCV overall carriage prevalence in a given country.

Given a fixed pre-PCV total pneumococcal carriage, increasing the initial proportion of VT among colonizing serotypes (quantity *F*, see Methods) by changing initial VT:NVT:Co-carriers ratio slowed elimination slightly (Figure 6C, Supplementary Figure 6), while maintaining *F* led to constant time-to-elimination (Supplementary Figure 7). With constant *F*, we further considered a range of competition levels (*k*_*V*_=*k*_*N*_=0.1, 0.25, 0.75) and found that time-to-elimination remained similar (time-to-elimination range: 4.3–6.3 years, 4.1–6.1 years, 3.6–5.8 years) (Supplementary Figure 8).

We changed the age-specific susceptibility parameter *β*^(*i*)^ by ±20% to simulate for high and low susceptibility. As expected, the total pneumococcal carriage pre-PCV increased with population susceptibility. However, the predicted effect of this parameter was moderate: transitioning from low to high population susceptibility resulted in 0.6–1.8 years longer time-to-elimination. This result may be explained by the fact that, for higher total carriage, more circulating VT had to be replaced. Considering the same population susceptibility level, countries with higher pre-PCV pneumococcal carriage had longer time-to-elimination, except for Canada and South Africa (Figure 6D), for which the contact rate and assortativity in children under 5 was the highest among the subset of 7 countries (Figures 5A, B).

In summary, of all the parameters tested, the vaccine parameters had the strongest impact on time-to-elimination, while the other parameters had a more moderate effect. This result highlights the need for accurate estimates of PCVs properties to predict the time scale of VT elimination in a target population.

## Discussion

The main goal of this study was to assess the effect of social contact structure on the impact of PCVs. To do so, we designed a pneumococcal transmission model, parameterized based on empirical data, and verified it against the observed decline in VT carriers among children in France, Alaska (US), Massachusetts (US), and the UK. Using the best available social contact matrices from 34 countries, our study showed that heterogeneity in contact structure alone can lead to a range of time-to-elimination and thus sensitively affect the impact of PCVs. In addition, they highlight the key role of contact features in children under 5 in VT elimination and provide new insights into the mechanisms of VT elimination. More broadly, these findings identify social contact structure as a new key variable affecting vaccine impact, with potential implications beyond PCVs.

Our model predicted a range of time-to-elimination (3.8–6 years) that is consistent with the literature [27–29], in support of the WHO’s recommendation that 5 years of post-PCV data are necessary to assess pneumococcal serotype replacement [30]. The modeled time-to-elimination in our study was the shortest among vaccinated age groups, who had the most social contacts, reflecting combined direct and indirect effectiveness, in contrast to age 0, who benefited from indirect effectiveness only (Figure 4). This finding aligns with the reported direct and indirect effects of PCV on carriage [11].

Different features of a contact matrix can have different effects on infectious disease dynamics. For example, assortativity, a well-studied feature measuring the extent of preferential mixing of individuals within the same demographic stratum, was shown to drive the spread of HIV infections differently in groups with different risks [31]. Another widely used feature is the number of social contacts, which was suggested as the main factor that explained the higher COVID rates among older adults in Italy [25]. We investigated these two features in our study and observed that the total contact rates in all countries followed a similar trend, with lower contact rates in extreme ages and a peak around school-age and teenage years (5–19y); contrastingly, there was a much higher variability in assortativity across countries. We found both features in children under 5 to be significant predictors for shorter time-to-elimination, indicating children under 5 as the key age group in driving the serotype replacement dynamics, despite having a lower contact rate than other age groups. The high contact rate and assortativity in children under 5 in Canada and South Africa could explain why they were the outliers with shorter time-to-elimination despite higher pre-PCV pneumococcal carriage (Figure 6D). These findings echo the evidence that children under 5 are key for pneumococcal transmission [32].

Intuitively, higher total contact rates would speed up the transmission dynamics and shorten time-to-elimination. The effect of assortativity can be explained by the high carriage prevalence in this age group. Children under 5 had the highest carriage prevalence, so a more assortative contact in this age group would promote the within-group transmission. In general, infection spreads faster for a high-risk group with assortative mixing because contacts with low-risk groups slow down the transmission dynamic [31]. These findings point to the vital role of contact patterns in the high-prevalence groups in infection transmission and can be the basis of infection preventive strategies.

In addition to social contact structure, we found vaccine factors to be the most influential parameters in the serotype replacement dynamics. This finding is consistent with the epidemiological evidence that locations with high vaccine coverage saw rapid carriage replacement [27]. We also found that rapid waning led to longer time-to-elimination. Furthermore, initial VT:NVT:Co-carriers ratio and population susceptibility had slight to moderate effects. Given the same overall carriage, initial VT:NVT:Co-carriers ratio only affected the time-to-replacement if the proportion of VT among circulating serotypes, *F*, was changed: higher *F* led to a longer time-to-elimination. The time-to-elimination was also longer in a more susceptible population whose overall carriage is higher. These results demonstrated that when the circulating VT burden is higher, it takes longer for replacement to be complete.

Our study has several limitations. When simulating the VT-carrier prevalence in children using location-specific parameter sets, our model captured the patterns in the observed data in the UK, Alaska (US), Massachusetts (US), but, with some discrepancy in the initial post-PCV era, in France. This discrepancy potentially stemmed from the partial uptake of PCV in the private market, reaching a vaccine coverage of over 20% in the year before vaccine introduction in our simulation [33]. In the simulations, we considered the uncertainty in vaccine efficacy but not in other parameters, which may have contributed to the VT-carrier prevalence decline being slightly slower than observed. For example, competition between VT and NVT could have enhanced the population-level impact of PCV [34]. Specifically, competition can be driven by direct competition between VT and NVT in the nasopharynx, or indirect competition due to innate and adaptive immunity that is cross-reactive for NVT and VT, or both [34,35]. We considered only direct competition in our model. We also did not consider seasonal fluctuations in contact rates and assortativity, which could affect the transmission of infections [36,37]. How this biased the estimated time-to-elimination depends on whether holidays increase the total contacts considering the change in both inter-age and intra-age contacts. Most of the contact matrices used in this study came from high-income countries, limiting our findings’ generalizability to other settings. In settings with high residual transmission despite persistently high vaccine coverage [38] in which the prevalence of underlying vulnerable groups may be important [39]. While there were published contact matrices for more countries [40,41], the ones used in our study offer the best age resolution to date. The variation of contact patterns across geographic locations and income settings is expected to be larger than observed in this study, and including them in future studies can help elucidate the phenomenon observed outside high-income settings. For instance, in high transmission settings, the pneumococcal reservoir may involve a wider age group, including school-age children [32]. Given the evidence of a higher extent of serotype replacement in indigenous children in Fiji [42] and in rural areas in Nigeria [43], future studies should explore further how contact patterns in sub-populations within the same country influence serotype replacement.

Despite these limitations, our study demonstrated how to combine contact matrices and mathematical modeling to unravel the dynamics between the host, the pathogen, and a public health intervention. The strengths of our study included using a neutral model, which is a parsimonious way to achieve initial levels of VT and NVT co-existence without specifying serotype-specific parameters, and using contact matrices of high age resolution, which allowed us to differentiate the transmission dynamics in every year of age. In addition, for parameters with variable estimates, we based our assumptions on non-linear models fitted to extracted data from observational studies. In conclusion, our findings demonstrate that, as for other vaccine-preventable diseases, social contact structure is a critical element for understanding the vaccine epidemiology of pneumococcus. Hence, we propose this element should be considered in future studies assessing the impact of PCVs and, more broadly, of other vaccines.

## Methods

We proceeded in three steps. First, we developed a dynamic model of pneumococcal carriage transmission (Figure 1, Supplementary Table 1). The parameter values were based on empirical data, taken from literature, or assumed (Table 2). We verified this model’s adequacy against the observed prevalence of VT carriers among children in France, Alaska (US), Massachusetts (US), and the UK (Table 1). Second, we simulated the dynamics of pneumococcal transmission after PCV introduction using contact matrices from 34 countries [16] and assessed the impact of social contact patterns on the dynamics of VT carriage decline. Third, we changed one key parameter (such as vaccine efficacy and population susceptibility) at a time in the simulations using a subset of 7 out of 34 contact matrices to investigate the effect of each key parameter on VT elimination. We describe the *Data*, the *Model*, and the details of these three steps in the *Analyses* below.

### Data

#### a) Contact matrices

We used the inferred contact matrices *M*_*ij*_ from [16]. The contacts, stratified by age yearly from 0 to 84, were derived from synthetic networks built using population census data and socio-demographic surveys in various settings, namely, household, school, workplace, and community. The overall contact matrix for a location is a weighted sum of the setting-specific contact matrices. *M*_*ij*_ gives the total number of daily contacts between age groups *i* and *j* per person in age group *i*, we applied reciprocity correction on *M*_*ij*_ and transformed it into *m*1_*ij*_, which gives the total annual contacts between age groups *i* and *j* per person in age group *i* and per person in age group *j* (density scale, as defined in [44]; see Supplementary Figure 2). To elucidate the effect of social contact patterns, we used a common population structure for all contact matrices in our simulations.

#### b) Carriage duration and prevalence

We extracted data about age-specific carriage duration and carriage prevalence from published studies identified through a scoping literature search (Supplementary Data 1, Supplementary Data 2).

Among the identified culture-based studies, we included the studies that reported median duration (n=8), because the duration of carriage has a left-skewed distribution, with few individuals showing lasting carriage. For the model of carriage duration with age, we used non-linear least square regression to estimate the parameters in the equation (Supplementary Figure 1):

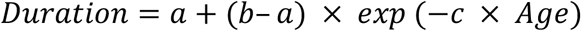

where *a* = 21 (standard error: 5.8), *b* = 62 (14.6), and *c* = 0.45 (0.4).

In the main analysis, we fixed the age-specific initial carriage prevalence 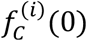 based on [26] and the age-specific susceptibility parameter *β*^(*i*)^ based on [47]. As sensitivity analyses, we used two other distributions of *β*^(*i*)^ over age, considering a lower carriage prevalence in age 0 and a higher carriage prevalence in all ages, to reflect the observed data from the identified culture-based pre-PCV carriage prevalence studies (n=17) (Supplementary Figure 4). After calibrating *β*^(*i*)^ for the assumed carriage prevalences, we re-simulated the time-to-elimination for all countries.

#### c) VT-carrier prevalence in children in the real world

We extracted the VT-carrier prevalence in children from the pre-to post-PCV era in 4 locations

- France [20], UK [21], Alaska, US [22], and Massachusetts, US [23] (Supplementary Data 3) to verify our model’s ability to reproduce the decline in VT-carrier prevalence following PCV introduction. The observed data come from cross-sectional surveys among children attending daycare centers or primary care clinics. In all four included studies, the detection of *S*.*pneumoniae* was culture-based and the serotyping was either by traditional Quellung reaction or molecular methods. For point estimates of carriage reported without uncertainty, we calculated the standard error (SE) for proportion and indicated the uncertainty limits as 1.96×SE from the mean.

### Model

#### a) Structure

We formulated a deterministic model that simulates the transmission of VT and NVT carriage based on the neutral null model proposed by [17]. Assuming a stable population (i.e., birth rate = death rate), susceptible individuals (*S*) become VT-carriers (*C*_*V*_) at the rate *λ*_*V*_, or NVT-carriers (*C*_*N*_) at the rate *λ*_*N*_. Mono-carriers *C*_*V*_ (or *C*_*N*_) can be colonized by the other serotype at rate *k*_*N*_ × *λ*_*N*_ (or *k*_*V*_ × *λ*_*V*_) and become co-carriers (*C*_*VN*_), and co-carriers return to mono-carriers *C*_*V*_ (or *C*_*N*_) at rate *c* × *k*_*V*_ × *λ*_*V*_ (or *c* × *k*_*N*_ × *λ*_*N*_). We assumed the inter-serotype competition parameter, *k*, to be 0.5 in the main analysis and tested a range of values (0.1, 0.25, 0.75) based on published estimates [18,45,46]. When *k*_*V*_=0.5, VT is half as likely to colonize an individual already colonized by NVT. We further assumed this competition to be symmetrical (*k*_*V*_ = *k*_*N*_) to ensure neutrality at initiation. The parameter *c*, representing the fraction of co-carriers returning to *C*_*V*_ (or *C*_*N*_) upon re-infection with VT (or NVT), was fixed to 0.5 to ensure neutrality [47].

The vaccine was introduced at time *t*_*V*_ and had a coverage of *p*_*V*_. Therefore, *p*_*V*_ was zero before time *t*_*V*_ and equal to *p*_*V*_ starting from time *t*_*V*_.

In our age-structured model, individuals moved from one age to the next year of age at an aging rate *δ*_*i*_ =1 per year. The whole population of newborns was unvaccinated. As individuals moved from age 0 to age 1, a fraction (*p*_*V*_) of the population was vaccinated and partially protected from pneumococcal colonization (superscript “(*V*, 1)”). The rest (1− *p*_*V*_) of age 0 stayed unvaccinated as they reached age 1 (superscript “(*N*, 1)”).

For the dynamics of the vaccinated individuals, the rate of VT carriage acquisition *λ*_*V*_ was reduced by a factor *ϵ*_*V*_, where *ϵ*_*V*_ represents the vaccine efficacy against acquisition of VT carriage. Vaccine-conferred immunity was assumed to wane at a rate *α*_*V*_, so that *1*/*α*_*V*_ represents the average duration of vaccine protection.

The age-specific acquisition rate, *λ*^(*i*)^, depends on *β*^(*i*)^, the cumulative number of carriers in the contactee age groups, *CC*^(*j*)^, and the per capita contact matrix, *m*1_*ij*_. The carriage acquisition rates for VT and NVT were expressed as:

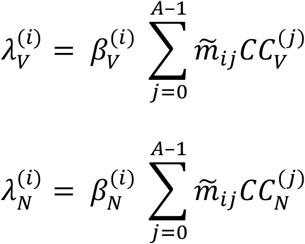

Where

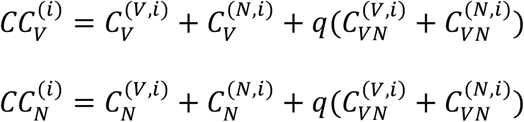

Here, *q* refers to the relative infectiousness with each serotype for co-carriers. Table 2 summarizes the parameters used in this study.

#### b) Outcome definition

In a neutral null model, one serotype is not assumed to have a fitness advantage over the other; therefore, co-carriers transmit either VT or NVT at equal probability. The relative infectiousness with each serotype for co-carriers, *q*, is set to 0.5, such that co-carriers are equally infectious as mono-carriers [17]. To ensure neutrality in the null model, we checked if

*F* was stable over time in the model without an effective vaccine, as suggested by [17] (Supplementary Figure 3). *F* is given by:

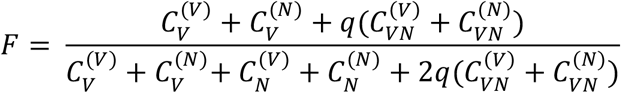

We defined time-to-elimination as the duration between vaccine introduction and the time when *F* dropped to 5% of its initial value in age 0, representing a fully unvaccinated population and reflecting the indirect effect of PCV introduction. As time-to-elimination in all ages were highly correlated in each country, the choice of age had a negligible effect on the analyses comparing countries.

### Analyses

#### a) Model assessment

To verify the model, we used location-specific parameter sets to simulate the VT-carrier prevalence in children from the pre-to post-PCV era in 4 locations (Table 1) and compared the simulated values to the observed ones qualitatively.

We calibrated the model for each location by estimating the parameters *β*^(*i*)^. First, we performed a global search on 1000 values between –10 and 10, corresponding to *β* values between 0 and 1 on the logit-transformed scale. The values were sampled using Sobol’s sequence [48], a quasi-random sampling method, to ensure the global parameter space was searched thoroughly. The global search sought a set of *β*^(*i*)^ that minimized the total squared difference in simulated versus observed pre-PCV VT-carrier prevalence on the logarithmic scale in all age groups. Here, the age groups were defined based on the observed prevalences as 0 y, 1–4 y, 5–17 y, 18–39 y, 40–59 y, and 60–84 y. The best five solutions from the global search were used as the starting value for a local search using the Subplex algorithm [49] until the total squared difference was minimized or could not be further reduced after a maximum of 1000 evaluations. Given the uncertainty around this parameter, we simulated the VT-carrier prevalence in children in each location using a range of vaccine efficacy against colonization [50].

#### b) Effect of contact features

To investigate the effect of social contact structure on the replacement dynamics, we first summarized the contact matrices using two age group-specific features – contact rate and assortativity – and then explored the relationship between these features and the time-to-elimination. Here, the age groups were defined as 0–4 y, 5–9 y, 10–19 y, 20–39 y, 40–59 y, and 60–84 y, to be consistent with the parameter value assignment (Table 2). We defined contact rate as the average total daily contacts in an age group and assortativity as the fraction of contacts from within the age group out of total contacts for each age in the age group.

We described the distribution of total contact and assortativity over age in all 34 contact matrices (Figures 5A, B) and explored the association between time-to-elimination and these contact features in all age groups (Supplementary Figure 9).

Based on the strong negative correlation between the two contact features and time-to-elimination portrayed in children under 5 (Figure 5C), we performed a regression analysis on time-to-elimination with standardized contact rate and standardized assortativity in this age group as predictors in a GLM with a log link (Figure 5D). We reported the effect estimates with 95%CI for both variables and assessed the goodness-of-fit with R-squared. We further tested the out-of-sample prediction performance of the GLM containing only these two predictors by leaving 4 contact matrices out as the test set and using the remaining 30 contact matrices as the training set. We repeated this procedure 10 times and reported the MRAE for each iteration (Supplementary Table 2).

#### c) Effect of key parameters

To delineate the individual effect of the key parameters— vaccine efficacy, vaccine coverage, immunity waning, the initial proportions of VT and NVT carriers, and population susceptibility—on time to elimination, we varied them one at a time using a range of values and measured the time-to-elimination.

Vaccine efficacy and coverage were considered key parameters because these contributed to the selective pressure that drives serotype replacement. We varied vaccine efficacy between 33–77% based on the observed efficacy with uncertainty in a community randomized trial [50], consistent with the findings of a systematic review [51]. Other than no waning, we tested a range of durations of vaccine-conferred immunity, ranging from 3 to 10 years [52,53]. Evidence suggests pre-PCV serotype distribution in carriage and diseases as important predictors of vaccine impact [54]; therefore, we tested a range of initial proportions of VT-carriers (*f*_*V*_(0)), NVT-carriers (*f*_*N*_(0)), and implicitly, co-carriers (1–*f*_*V*_(0)–*f*_*N*_(0)), either allowing the proportion of VT among colonizing serotypes (*F*) to fluctuate or be fixed at 0.65. For constant *F*, we further considered a range of competition levels (*k*_*V*_=*k*_*N*_=0.1, 0.25, 0.75) in the sensitivity analysis.

In these model experiments, the age-specific overall carriage remained constant. Lastly, to investigate the dynamics under different population susceptibilities, we changed the age-specific susceptibility parameter, *β*^(*i*)^, by ±20% compared to the baseline value, which led to higher and lower overall carriage, respectively. In each simulation, we used 7 contact matrices from [16] to see if the effect of each key parameter differs by social contact structure.

##### Numerical implementation

All analyses were conducted in RStudio with R version 4.2.2 (R) and the non-linear model fitting was performed using the base package “stats” [55]. The transmission model was implemented using the package “pomp” version 4.6 [56]. All optimization procedures were implemented using the algorithms available in the package “nloptr” version 2.0.3 [57].

## Data Availability

The code and data are available online at Edmond, the Open Data Repository from the Max Planck Society.

https://doi.org/10.17617/3.RIGYAK

## Code and data availability

The code and data are available from Edmond, the Open Data Repository from the Max Planck Society: https://doi.org/10.17617/3.RIGYAK.

