## Supplementary_materials for "Assessing the effect of social contact structure on the impact of pneumococcal conjugate vaccines"


| | | |
|---|---|---|
| Figure 1 | Modelled vs. observed duration of carriage with increasing age | 1 |
| Figure 2 | Transforming the contact matrices from Mistry et al. 2021 | 2 |
| Figure 3 | Checking model assumption: neutral null model | 5 |
| Table 1 | Model structure: state variables | 6 |
| Figure 4 | Assumed vs. observed carriage prevalences and different susceptibility distributions over age | 9 |
| Figure 5 | Effect of changing vaccine efficacy and coverage on time-to-elimination | 11 |
| Figure 6 | Effect of initial proportions of VT, NVT and Co-carriers on time-to-elimination, with changing initial proportion of VT among all colonizing serotypes ($F$) | 12 |
| Figure 7 | Effect of initial proportions of VT, NVT and Co-carriers on time-to-elimination, with fixed initial proportion of VT among all colonizing serotypes ($F$) | 13 |
| Figure 8 | Effect of competition on time-to-elimination with fixed proportion of VT among all colonizing serotypes ($F$) | 14 |
| Figure 9 | Association of time-to-elimination and features of contact patterns in all age groups | 15 |
| Table 2 | Out-of-sample prediction using contact rate and assortativity as predictors | 17 |
| References | | 18 |

| | |
|---|---|
| Data 1 | Extracted data for fitting clearance rate (available as a data file) |
| Data 2 | Extracted data for checking assumption on carriage prevalence (available as a data file) |
| Data 3 | Extracted data for verifying simulated VT-carriage in children (available as a data file) |

**Figure 1. Modelled vs. observed duration of carriage with increasing age**

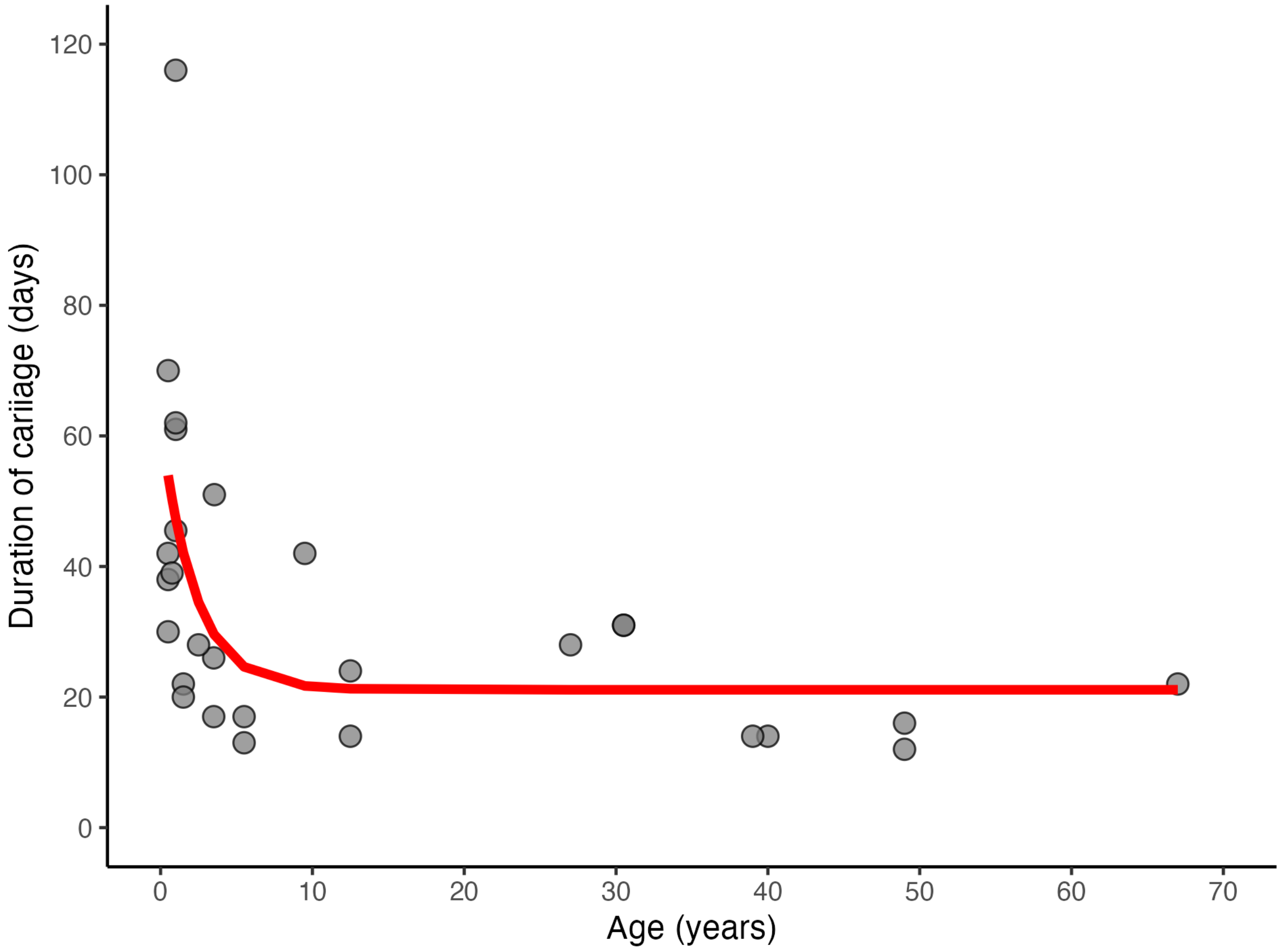


To obtain the duration of carriage, we fitted a non-linear function of age to the extracted duration of carriage from published longitudinal carriage studies identified through a scoping literature search (Data 1). We first searched for observational studies on pneumococcal carriage duration on PubMed and further identified relevant studies from the references of the initially included studies. We selected culture-based studies to allow the inclusion of the maximum number of studies because the majority of the early studies relied on culture-based detection. Further, because the duration of carriage has a left-skewed distribution, with few individuals showing lasting carriage, we preferred median to mean reporting and included studies that reported median duration. A total of 8 studies were included [1–8]. For the model of carriage duration with age, we used the non-linear least square algorithm to estimate the parameters in the equation:

$$Duration = a + (b - a) \times exp\ (-c \times Age)$$

where $a = 21$ (standard error: 5.8), $b = 62$ (14.6), and $c = 0.45$ (0.4).

Figure 1 shows the extracted data from observational studies as grey points and the modeled duration of carriage with a red curve.

**Figure 2. Transforming the contact matrices from Mistry et al. 2021**

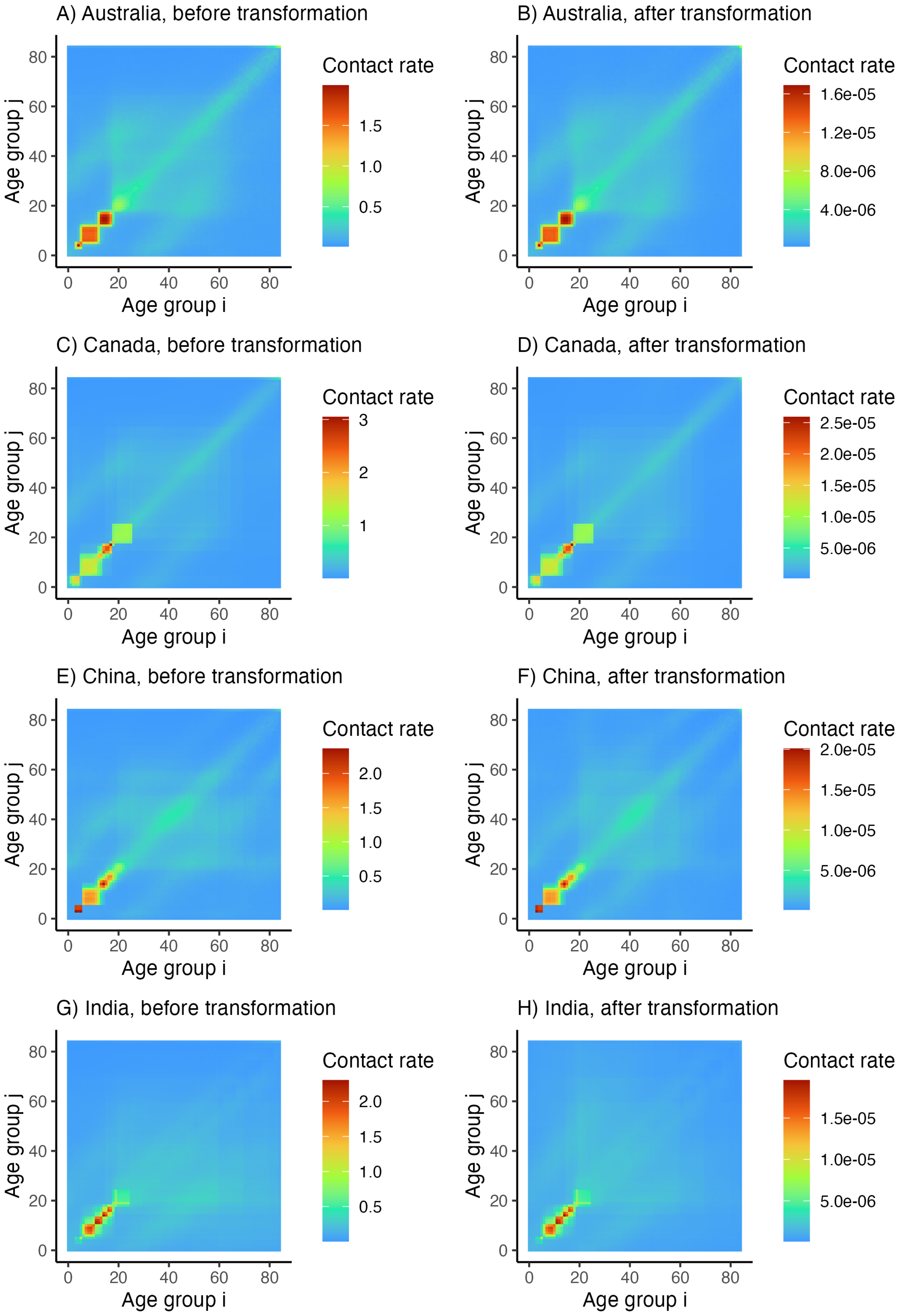

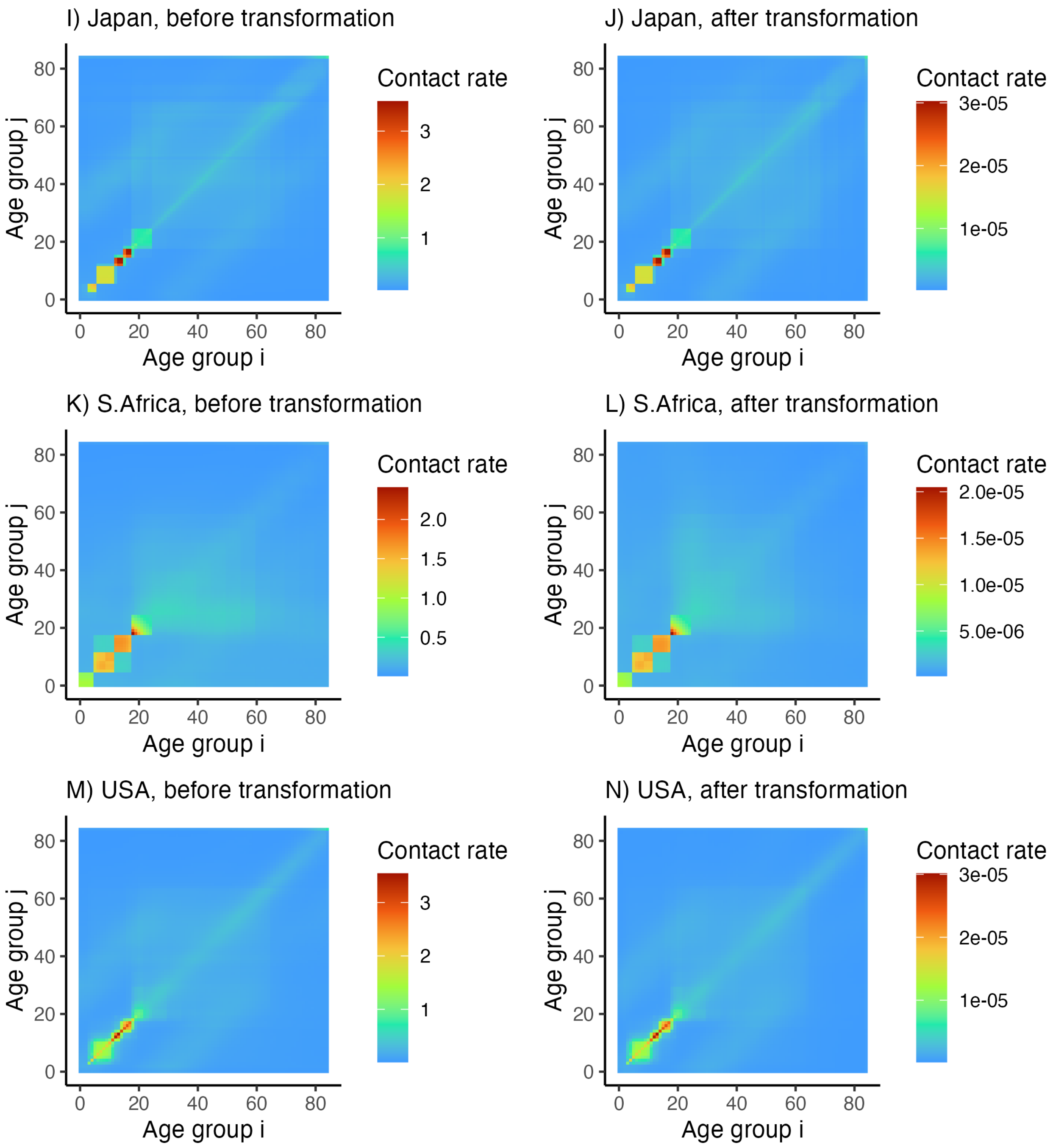


Let $M = (M_{ij})$ represent the original contact rate matrix calculated in [9]. By definition, $M_{ij} = \frac{E_{ij}}{N_i}$, where $E_{ij}$ represents the number of daily contacts between age groups $i$ and $j$, and $N_i$ the population size of age group $i$. Because of the necessary reciprocity of total contacts (i.e., $E_{ij} = E_{ji}$), the per capita contact matrix $m_{ij} = \frac{M_{ij}}{N_j} = \frac{E_{ij}}{N_i N_j}$ should be symmetric. To ensure this symmetry, one can calculate a contact matrix $\widetilde{M}$ corrected for reciprocity based on the population structure in the study population [10]:

$$\widetilde{M}_{ij} = \frac{1}{2N_i}(M_{ij}N_i + M_{ji}N_j)$$

As a result, the per capita contact matrix:

$$\widetilde{m}_{ij} = \frac{M_{ij}N_i + M_{ji}N_j}{2N_iN_j}$$

is symmetric, as it should be.

Finally, we multiplied the per capita matrix by 365 to obtain a per capita annual contact matrix because all rates were per year in the simulations.

Figure 2 (A–N) shows the pre- and post-transformation contact patterns of a subset of 7 out of the 34 contact matrices as examples.

**Figure 3. Checking model assumption: neutral null model**

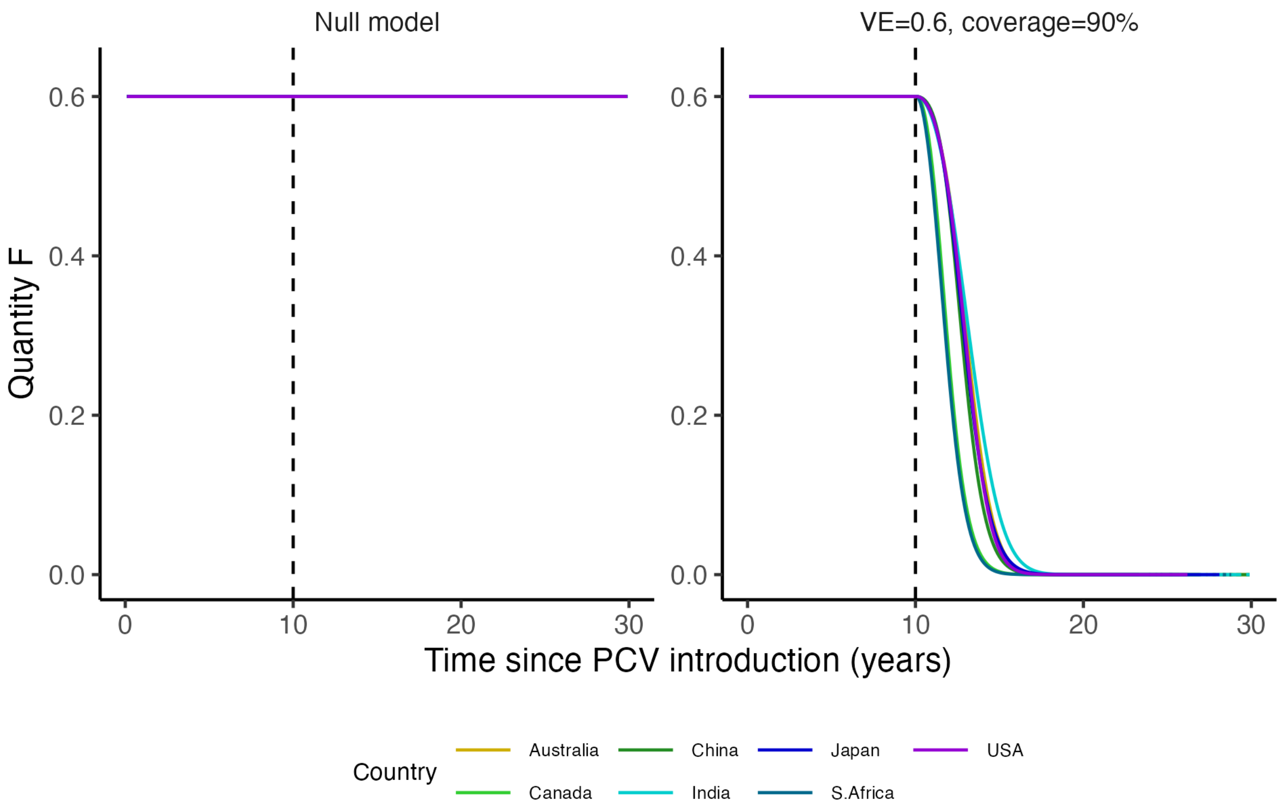


To check if our null model fulfils the neutrality criterion, we tracked proportion of VT among all colonizing serotypes ($F$) as proposed by [11]. The quantity $F$ is given by:

$$F = \frac{C_V^{(V)} + C_V^{(N)} + q(C_{VN}^{(V)} + C_{VN}^{(N)})}{C_V^{(V)} + C_V^{(N)} + C_N^{(V)} + C_N^{(N)} + 2q(C_{VN}^{(V)} + C_{VN}^{(N)})}$$

In a neutral null model, where one serotype is not assumed to have a fitness advantage over the other, any level of co-existence should be permitted and the proportions of the two serotypes should not converge to a global equilibrium (e.g., $F$ = 50%) without explicit mechanism [11]. In other words, neutral models do not lead to a 50-50 coexistence by default. Specifically, $F$ should remain constant regardless of the initial proportions of VT and NVT under no intervention, where the intervention represents a mechanism that gives one serotype a fitness advantage over the other.

The left panel of Figure 3 shows that $F$ remained stable when a null-impact vaccine was introduced for all contact matrices. The right panel shows that $F$ declined after the introduction of a vaccine with vaccine efficacy=0.6 at 90% coverage, at different rates in different contact structures.

**Table 1. Model structure: state variables**

| State variable | Definition |
|---|---|
| $S^{(V,i)}$ | Susceptible, vaccinated, age group $i$ |
| $C_V^{(V,i)}$ | Colonized by VT, vaccinated, age group $i$ |
| $C_N^{(V,i)}$ | Colonized by NVT, vaccinated, age group $i$ |
| $C_{VN}^{(V,i)}$ | Colonized by VT and NVT, vaccinated, age group $i$ |
| $S^{(N,i)}$ | Susceptible, unvaccinated, age group $i$ |
| $C_V^{(N,i)}$ | Colonized by VT, unvaccinated, age group $i$ |
| $C_N^{(N,i)}$ | Colonized by NVT, unvaccinated, age group $i$ |
| $C_{VN}^{(V,i)}$ | Colonized by VT and NVT, unvaccinated, age group $i$ |

The transmission dynamic is described by the following system of ordinary differential equations:

**Equations in (unvaccinated) newborns ($i = 0$)**

Newborns are assumed to be non-carriers and, therefore, directly enter the $S^{(N,0)}$ compartment.

$\delta_0$ represents the aging rate, equal to $\frac{1}{a_0}$, where $a_0$ represents the age span of age group 0.

$N = \sum_{i=0}^{A-1} N_i$ is the total population size (summed across all age groups).

$\mu$ is the per capita birth rate. To keep the population constant, this is calculated as $\mu^{-1} = a = \sum_i a_i$, where $a_i$ is the age span of age groups $i$ and $a$ represents the assumed lifespan. For example, if one assumes a lifespan $a = 80$ years, then the birth rate equals $\mu = a^{-1} = \frac{1}{80}$ per year.

In the model, newborns are born at the rate $\mu N$, then individuals age across the age groups, and all die exactly at the age of 84. This simplified demographic model is known as type-I mortality distribution [12].

$$\frac{dS^{(V,0)}}{dt} = 0$$

$$\frac{dC_V^{(V,0)}}{dt} = 0$$

$$\frac{dC_N^{(V,0)}}{dt} = 0$$

$$\frac{dC_{VN}^{(V,0)}}{dt} = 0$$

$$\frac{dS^{(N,0)}}{dt} = \mu N - (\lambda_V^{(0)} + \lambda_N^{(0)})S^{(N,0)} + \gamma_0(C_V^{(N,0)} + C_N^{(N,0)} + C_{VN}^{(N,0)}) - \delta_0 S^{(N,0)}$$

$$\frac{dC_V^{(N,0)}}{dt} = \lambda_V^{(0)}(S^{(N,0)} + c\, k_V C_{VN}^{(N,0)}) - k_N\lambda_N^{(0)}C_V^{(N,0)} - (\gamma_0 + \delta_0)C_V^{(N,0)}$$

$$\frac{dC_N^{(N,0)}}{dt} = \lambda_N^{(0)}(S^{(N,0)} + c\, k_N C_{VN}^{(N,0)}) - k_V\lambda_V^{(0)}C_N^{(N,0)} - (\gamma_0 + \delta_0)C_N^{(N,0)}$$

$$\frac{dC_{VN}^{(N,0)}}{dt} = k_N\lambda_N^{(0)}C_V^{(N,0)} + k_V\lambda_V^{(0)}C_N^{(N,0)} - c(k_N\lambda_N^{(0)} + k_V\lambda_V^{(0)})C_{VN}^{(N,0)} - (\gamma_0 + \delta_0)C_{VN}^{(N,0)}$$

**Equations in infants receiving vaccination ($i = 1$)**

Write $v(t) = p_V \; for \; (t \geq t_V)$, the time-varying vaccine coverage, where $t_V$ represents the time point of PCV introduction and $pV$ the proportion of infants vaccinated after PCV introduction.

Vaccination is assumed to occur at age 0, that is, when the newborns age to the second age group. The other properties of the vaccine are its efficacy against acquisition of V-serotypes (denoted by $\epsilon_V$) and its rate of waning protection (denoted by $\alpha_V$, where $1/\alpha_V$ represents the average duration of vaccine protection).

$$\frac{dS^{(V,1)}}{dt} = v(t)\delta_0 S^{(N,0)} - [\lambda_V^{(1)}(1-\epsilon_V) + \lambda_N^{(1)}]S^{(V,1)} + \gamma_1(C_V^{(V,1)} + C_N^{(V,1)} + C_{VN}^{(V,1)}) - (\alpha_V + \delta_1)S^{(V,1)}$$

$$\frac{dC_V^{(V,1)}}{dt} = v(t)\delta_0 C_V^{(N,0)} + \lambda_V^{(1)}(1-\epsilon_V)(S^{(V,1)} + c\, k_V C_{VN}^{(V,1)}) - k_N\lambda_N^{(1)}C_V^{(V,1)} - (\alpha_V + \gamma_1 + \delta_1)C_V^{(V,1)}$$

$$\frac{dC_N^{(V,1)}}{dt} = v(t)\delta_0 C_N^{(N,0)} + \lambda_N^{(1)}(S^{(V,1)} + c\, k_N C_{VN}^{(V,1)}) - k_V\lambda_V^{(1)}(1-\epsilon_V)C_N^{(V,1)} - (\alpha_V + \gamma_1 + \delta_1)C_N^{(V,1)}$$

$$\frac{dC_{VN}^{(V,1)}}{dt} = v(t)\delta_0 C_{VN}^{(N,0)} + k_N\lambda_N^{(1)}C_V^{(V,1)} + k_V\lambda_V^{(1)}(1-\epsilon_V)C_N^{(V,1)} - c[k_N\lambda_N^{(1)} + k_V\lambda_V^{(1)}(1-\epsilon_V)]C_{VN}^{(V,1)} - (\alpha_V + \gamma_1 + \delta_1)C_{VN}^{(V,1)}$$

$$\frac{dS^{(N,1)}}{dt} = [1 - v(t)]\delta_0 S^{(N,0)} - (\lambda_V^{(1)} + \lambda_N^{(1)})S^{(N,1)} + \gamma_1\left(C_V^{(N,1)} + C_N^{(N,1)} + C_{VN}^{(N,1)}\right) + \alpha_V S^{(V,1)} - \delta_1 S^{(N,1)}$$

$$\frac{dC_V^{(N,1)}}{dt} = [1 - v(t)]\delta_0 C_V^{(N,0)} + \lambda_V^{(1)}(S^{(N,1)} + c\, k_V C_{VN}^{(N,1)}) - k_N\lambda_N^{(1)}C_V^{(N,1)} + \alpha_V C_V^{(V,1)} - (\gamma_1 + \delta_1)C_V^{(N,1)}$$

$$\frac{dC_N^{(N,1)}}{dt} = [1 - v(t)]\delta_0 C_N^{(N,0)} + \lambda_N^{(1)}(S^{(N,1)} + c\, k_N C_{VN}^{(N,1)}) - k_V\lambda_V^{(1)}C_N^{(N,1)} + \alpha_V C_N^{(V,1)} - (\gamma_1 + \delta_1)C_N^{(N,1)}$$

$$\frac{dC_{VN}^{(N,1)}}{dt} = [1 - v(t)]\delta_0 C_{VN}^{(N,0)} + k_N\lambda_N^{(1)}C_V^{(N,1)} + k_V\lambda_V^{(1)}C_N^{(N,1)} - c(k_N\lambda_N^{(1)} + k_V\lambda_V^{(1)})C_{VN}^{(N,1)} + \alpha_V C_{VN}^{(V,1)} - (\gamma_1 + \delta_1)C_{VN}^{(N,1)}$$

**Equations in older age groups ($i = 2, \ldots, A-1$)**

In the older age groups, no more vaccination is assumed, and the dynamic is described by the following system of ordinary differential equations:

$$\frac{dS^{(V,i)}}{dt} = \delta_{i-1}S^{(V,i-1)} - [\lambda_V^{(i)}(1-\epsilon_V) + \lambda_N^{(i)}]S^{(V,i)} + \gamma_i(C_V^{(V,i)} + C_N^{(V,i)} + C_{VN}^{(V,i)}) - (\alpha_V + \delta_i)S^{(V,i)}$$

$$\frac{dC_V^{(V,i)}}{dt} = \delta_{i-1}C_V^{(V,i-1)} + \lambda_V^{(i)}(1-\epsilon_V)(S^{(V,i)} + c\, k_V C_{VN}^{(V,i)}) - k_N\lambda_N^{(i)}C_V^{(V,i)} - (\alpha_V + \gamma_i + \delta_i)C_V^{(V,i)}$$

$$\frac{dC_N^{(V,i)}}{dt} = \delta_{i-1}C_N^{(V,i-1)} + \lambda_N^{(i)}(S^{(V,i)} + c\,k_N C_{VN}^{(V,i)}) - k_V\lambda_V^{(i)}(1-\epsilon_V)C_N^{(V,i)} - (\alpha_V + \gamma_i + \delta_i)C_N^{(V,i)}$$

$$\frac{dC_{VN}^{(V,i)}}{dt} = \delta_{i-1}C_{VN}^{(V,i-1)} + k_N\lambda_N^{(i)}C_V^{(V,i)} + k_V\lambda_V^{(i)}(1-\epsilon_V)C_N^{(V,i)} - c[k_N\lambda_N^{(i)} + k_V\lambda_V^{(i)}(1-\epsilon_V)]C_{VN}^{(V,i)} - (\alpha_V + \gamma_i + \delta_i)C_{VN}^{(V,i)}$$

$$\frac{dS^{(N,i)}}{dt} = \delta_{i-1}S^{(N,i-1)} - (\lambda_V^{(i)} + \lambda_N^{(i)})S^{(N,i)} + \gamma_i\left(C_V^{(N,i)} + C_N^{(N,i)} + C_{VN}^{(N,i)}\right) + \alpha_V S^{(V,i)} - \delta_i S^{(N,i)}$$

$$\frac{dC_V^{(N,i)}}{dt} = \delta_{i-1}C_V^{(N,i-1)} + \lambda_V^{(i)}(S^{(N,i)} + c\,k_V C_{VN}^{(N,i)}) - k_N\lambda_N^{(i)}C_V^{(N,i)} + \alpha_V C_V^{(V,i)} - (\gamma_i + \delta_i)C_V^{(N,i)}$$

$$\frac{dC_N^{(N,i)}}{dt} = \delta_{i-1}C_N^{(N,i-1)} + \lambda_N^{(i)}(S^{(N,i)} + c\,k_N C_{VN}^{(N,i)}) - k_V\lambda_V^{(i)}C_N^{(N,i)} + \alpha_V C_N^{(V,i)} - (\gamma_i + \delta_i)C_N^{(N,i)}$$

$$\frac{dC_{VN}^{(N,i)}}{dt} = \delta_{i-1}C_{VN}^{(N,i-1)} + k_N\lambda_N^{(i)}C_V^{(N,i)} + k_V\lambda_V^{(i)}C_N^{(N,i)} - c(k_N\lambda_N^{(i)} + k_V\lambda_V^{(i)})C_{VN}^{(N,i)} + \alpha_V C_{VN}^{(V,i)} - (\gamma_i + \delta_i)C_{VN}^{(N,i)}$$

**Initial conditions**

The model was first run to simulate the pre-vaccine era, so all corresponding initial conditions were set to $0 = (X^{(V,i)} = 0)$ . For the initial conditions in the non-vaccinated groups, we defined the following parameters:

$f_C^{(i)}(0)$ : initial prevalence of carriers in age group $i$
$f_V(0)$ : proportion of VT carriers / all carriers
$f_N(0)$ : proportion of NVT carriers / all carriers
$f_{VN}(0) = 1 - f_V(0) - f_N(0)$ : proportion of dual carriers/all carriers

With these parameters, the state variables were initialized as follows:

$$S^{(N,i)}(0) = N_i \times [1 - f_C^{(i)}(0)]$$
$$C_V^{(N,i)}(0) = N_i \times f_C^{(i)}(0) \times f_V(0)$$
$$C_N^{(N,i)}(0) = N_i \times f_C^{(i)}(0) \times f_N(0)$$
$$C_{VN}^{(N,i)}(0) = N_i \times f_C^{(i)}(0) \times [1 - f_V(0) - f_N(0)]$$

**Figure 4. Assumed vs. observed carriage prevalences and different susceptibility distributions over age**

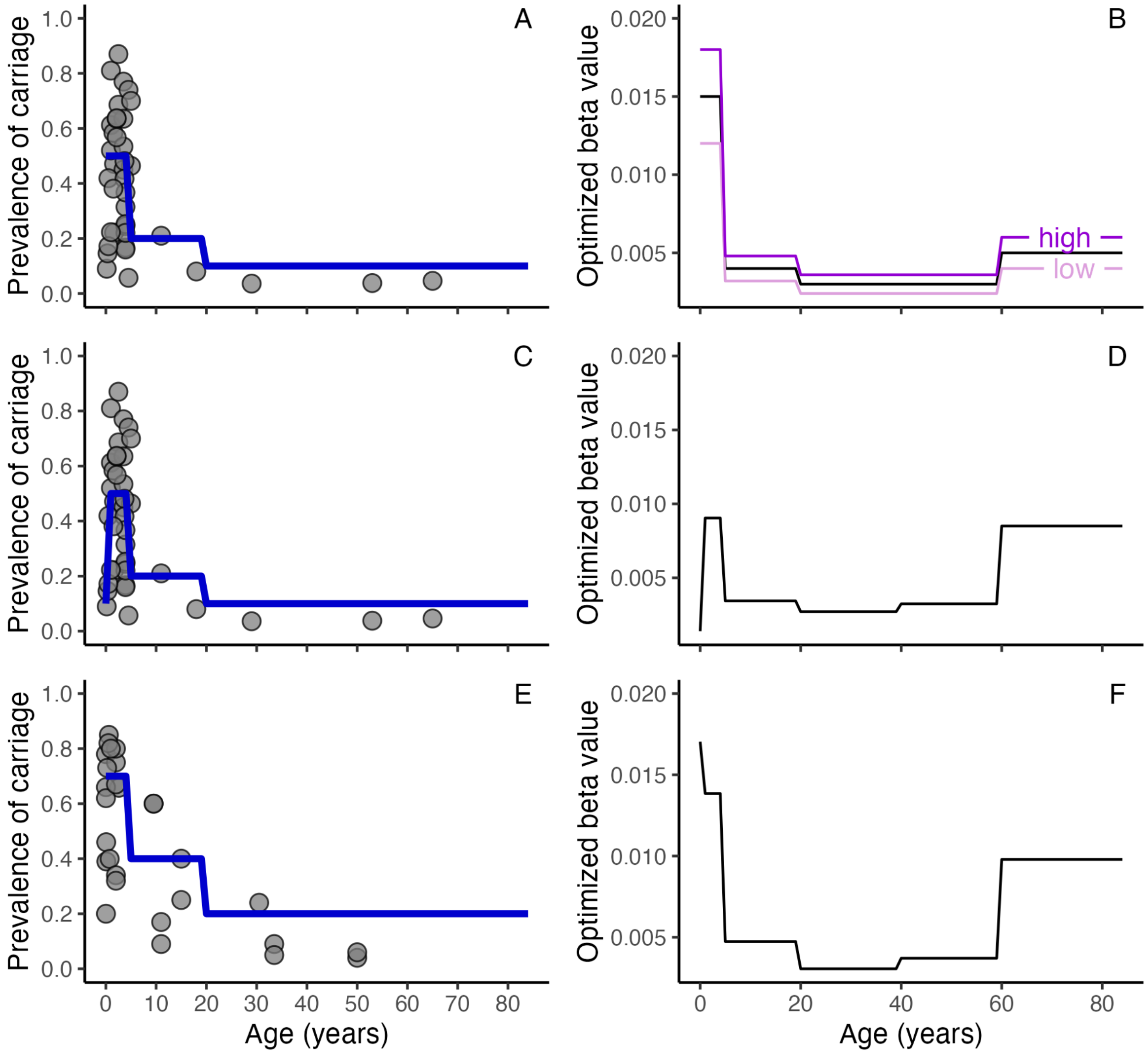


In Figure 4 left panels, the points show the observed carriage prevalence by age extracted from published observational studies (Data 2) conducted in high-income countries (HIC) (panels A, C) and low-income countries (LMIC) (panel E); the blue lines mark the assumed initial prevalence of carriers in the main analysis (panel A) and the sensitivity analyses (panels C, E). The right panels show the age-specific susceptibility parameter values used in the main analysis (panel B) and the sensitivity analyses (panels D, F).

In the main analysis, we assumed the initial overall carriage prevalence to be 50% in age 0-4, 20% in age 5-19, and 10% in age 20-84, based on a review [13], reflecting the carriage prevalence in the settings of high-income countries (panel A). We fixed the age-specific susceptibility parameter $\beta^{(i)}$ for low, medium, and high population susceptibility (panel B) when quantifying the influence of this parameter to time-to-elimination (results in main text Figure 6D).

As sensitivity analyses, we used different susceptibility distributions over age considering two scenarios based on a scoping literature review.

We searched for observational studies on pneumococcal carriage prevalence on PubMed and further identified relevant studies from the references of the initially included studies. We included culture-based studies conducted in the pre-PCV era that reported either overall (including both VT and NVT) or VT carriage prevalence because VT-carriers accounted for most of the carriage in the pre-PCV era. After excluding publications that were based on the same sample, a total of 17 studies were included (HIC: n=11 [14–24], LMIC: n=6 [1,3,4,6,25,26]). We observed that the carriage prevalence at age 0 was lower than in children aged 1–4, and that carriage prevalence was higher at all ages in LMIC compared with HIC. The income group of countries were based on the World Development Indicators 2008 [27].

Based on these observations, we considered a lower prevalence of carriers at age 0 (10% instead of 50%) due to the time lag from birth to first pneumococcal acquisition (panel C) and optimized $\beta^{(i)}$ (panel D). The resulted time-to-elimination ranged from 4.2–7.1 years.

We then considered higher carriage prevalences for all ages (70% in age 0–4, 40% in age 5–19, and 20% in age 20–84) to mimic settings with higher pneumococcal burden (panel E), which is commonly observed in low-income countries (LIC) [13,28], and optimized $\beta^{(i)}$ (panel F). The resulted time-to-elimination ranged from 4.4–6.9 years.

**Figure 5. Effect of changing vaccine efficacy and coverage on time-to-elimination**

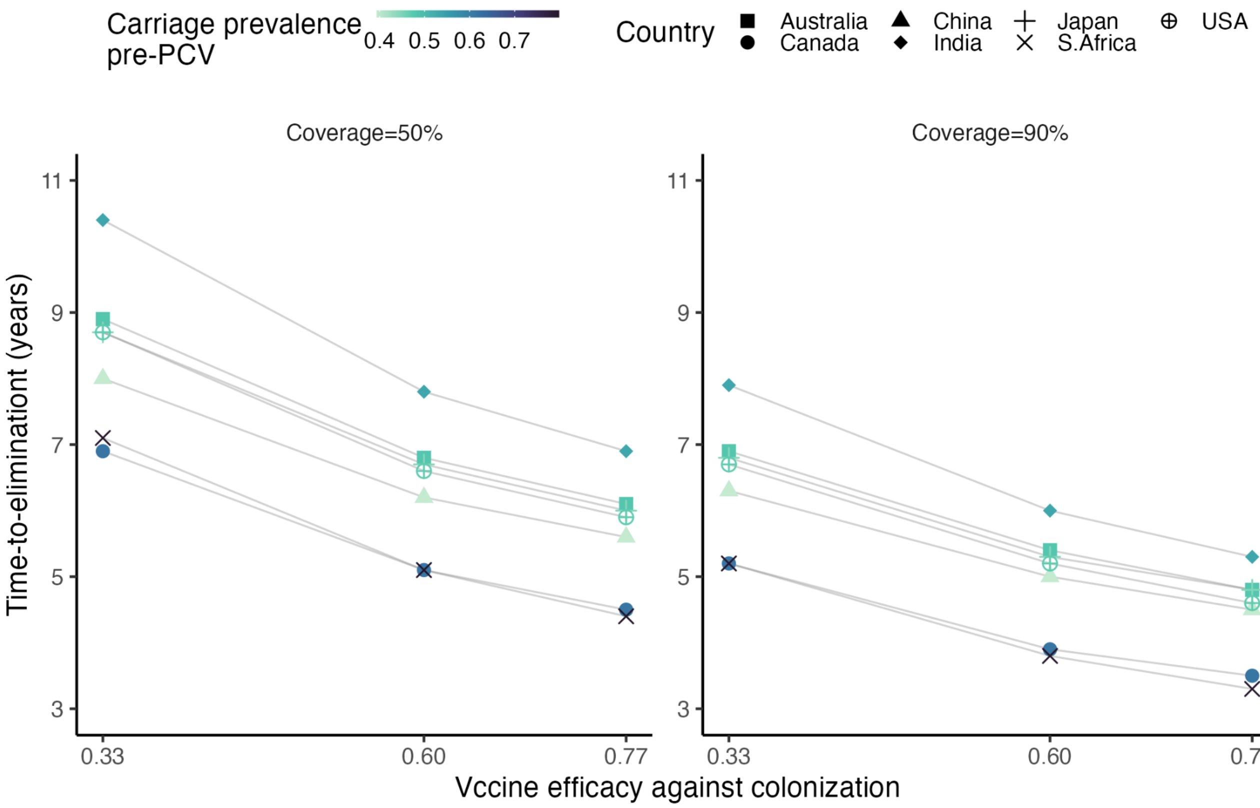


The change in time-to-elimination due to vaccine efficacy was larger when vaccine coverage was lower. At 50% coverage, switching from a less efficacious vaccine (vaccine efficacy=33%) to a highly efficacious vaccine (77%) resulted in 2.5–3.5 years shorter time-to-elimination (left panel). When vaccine coverage reached 90%, using a highly efficacious vaccine led to a 1.9–2.6-year reduction in time-to-elimination compared with a less efficacious vaccine (right panel, same as main text Figure 6A).

**Figure 6. Effect of initial proportions of VT, NVT and Co-carriers on time-to-elimination, with changing initial proportion of VT among all colonizing serotypes ($F$)**

| $f_V(0)$ | $f_N(0)$ | $f_{Co}(0)$ | $F$ |
|---|---|---|---|
| 0.2 | 0.2 | 0.6 | 0.5 |
| 0.4 | 0.2 | 0.4 | 0.6 |
| 0.4 | 0.4 | 0.2 | 0.5 |
| 0.6 | 0.2 | 0.2 | 0.7 |
| 0.6 | 0.4 | 0 | 0.6 |
| 0.8 | 0.2 | 0 | 0.8 |

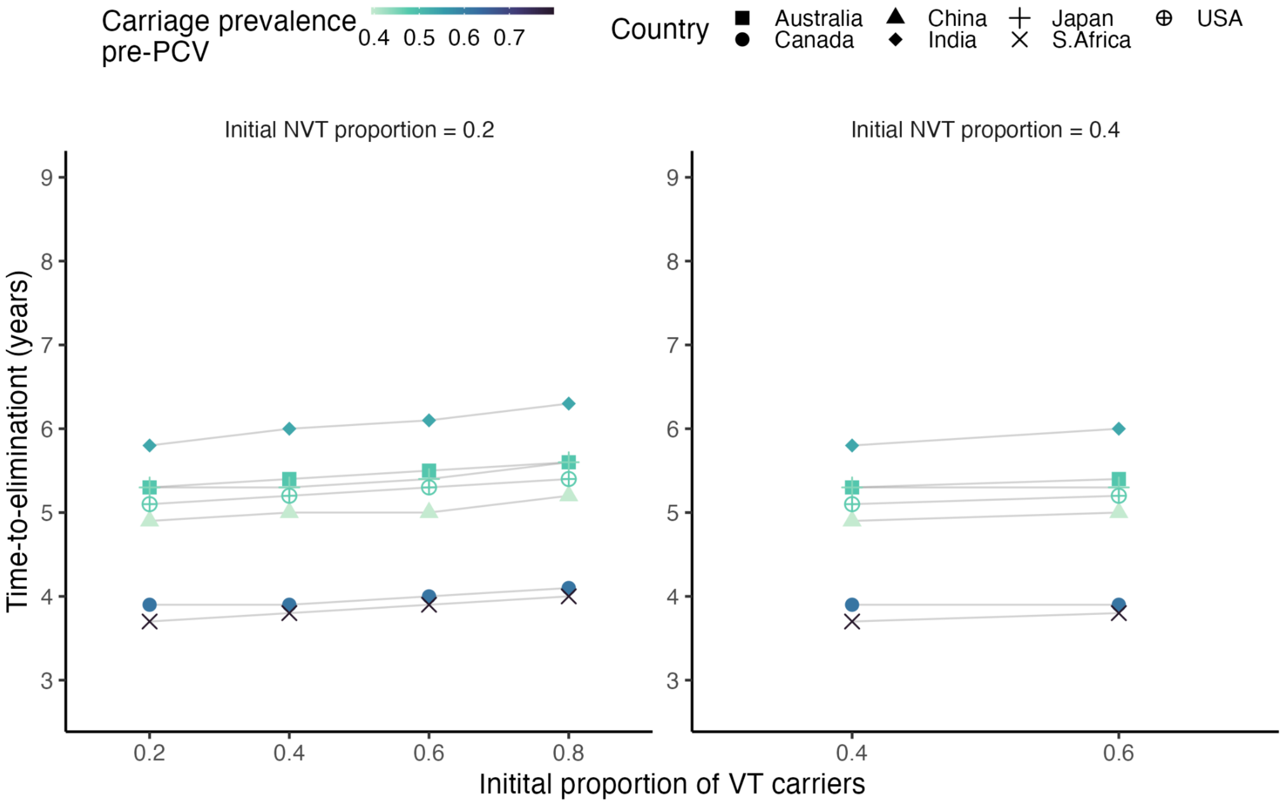


We varied the initial proportion of VT-carriers ($f_V(0)$) between 0.2 and 0.8, and the initial proportion of NVT-carriers ($f_N(0)$) between 0.2 and 0.4, with the proportion of co-carriers ($f_{Co}(0)$) implicitly determined ($1 - f_V(0) - f_N(0)$). The combinations of initial VT:NVT:Co-carriers ratio were restricted to two conditions: (1) $f_V(0) + f_N(0) \leq 1$ because the sum of proportions cannot exceed one , and (2) $f_V(0) \geq f_N(0)$ because VT-carriers tended to be more prevalent than NVT carriers in the pre-PCV era (main text Table 1).

When $f_N(0)$ was fixed at 0.2, increasing $f_V(0)$ from 0.2 to 0.8 resulted in an increase in the initial proportion of VT among all colonizing serotypes ($F$) from 0.5 to 0.8, leading to a slightly longer time-to-elimination (left panel, same as main text Figure 6C).

This effect remained when $f_N(0)$ was fixed at 0.4 and $f_V(0)$ increased from 0.4 to 0.6, which resulted in an increase in initial $F$ from 0.5 to 0.6.

**Figure 7. Effect of initial proportions of VT, NVT and Co-carriers on time-to-elimination, with fixed proportion of VT among all colonizing serotypes (*F*)**

| $f_V(0)$ | $f_N(0)$ | $f_{Co}(0)$ | $F$ |
|---|---|---|---|
| 0.60 | 0.30 | 0.1 | 0.65 |
| 0.65 | 0.35 | 0 | 0.65 |
| 0.50 | 0.20 | 0.3 | 0.65 |
| 0.40 | 0.10 | 0.5 | 0.65 |

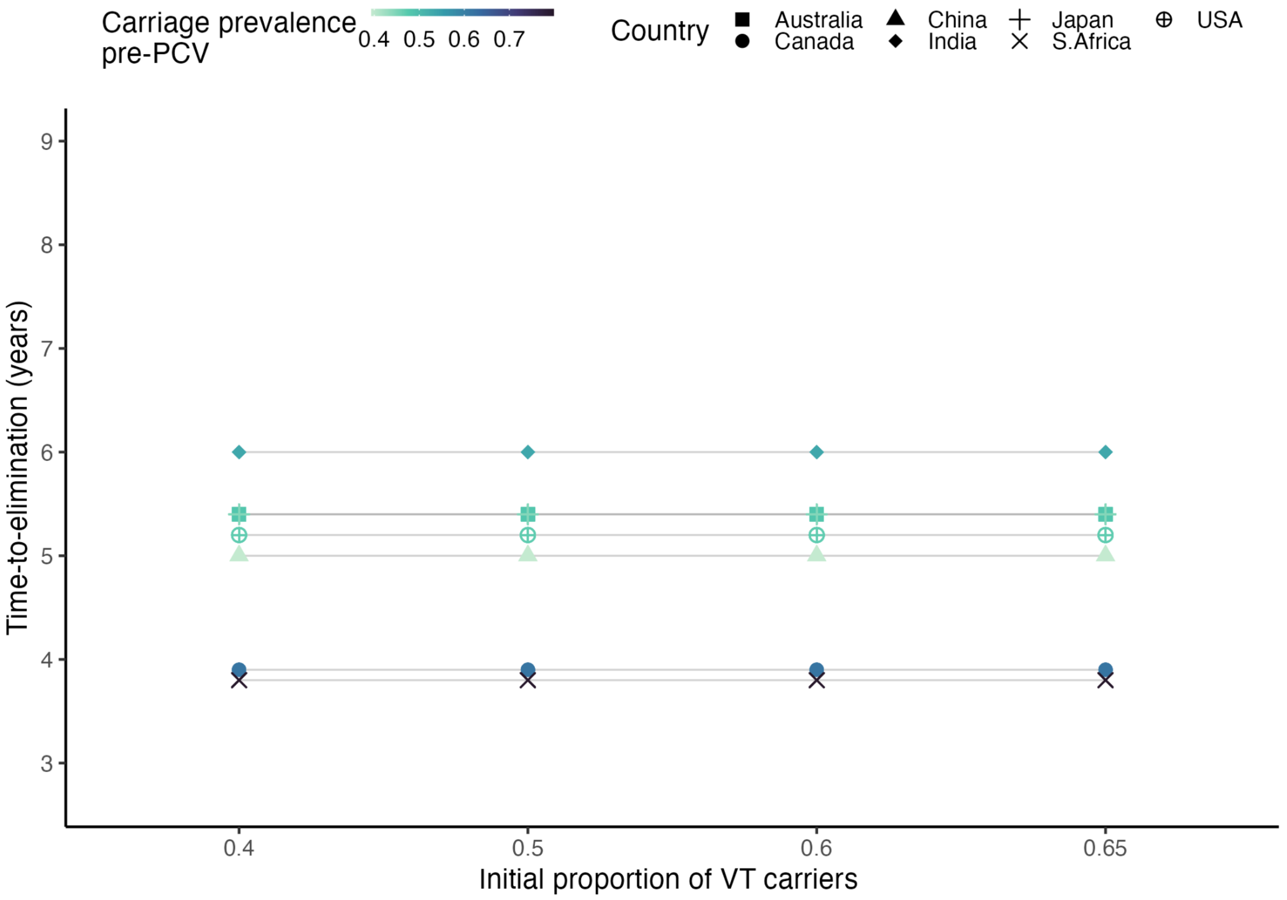


We varied the initial proportion of VT-carriers ($f_V(0)$), of NVT-carriers ($f_N(0)$), and implicitly, the initial proportion of co-carriers ($1 - f_V(0) - f_N(0)$), such that the initial proportion of VT among all colonizing serotypes ($F$) remained fixed at 0.65. The combinations of initial VT:NVT:Co-carriers ratio were restricted to two conditions: (1) $f_V(0) + f_N(0) \leq 1$ because the sum of proportions cannot exceed one , and (2) $f_V(0) \geq f_N(0)$ because VT-carriers tended to be more prevalent than NVT carriers in the pre-PCV era (main text Table 1).

With initial $F$ fixed at 0.65, the time-to-elimination remained constant regardless of the initial VT:NVT:Co-carriers ratio.

**Figure 8. Effect of competition on time-to-elimination with fixed proportion of VT among all colonizing serotypes (*F*)**

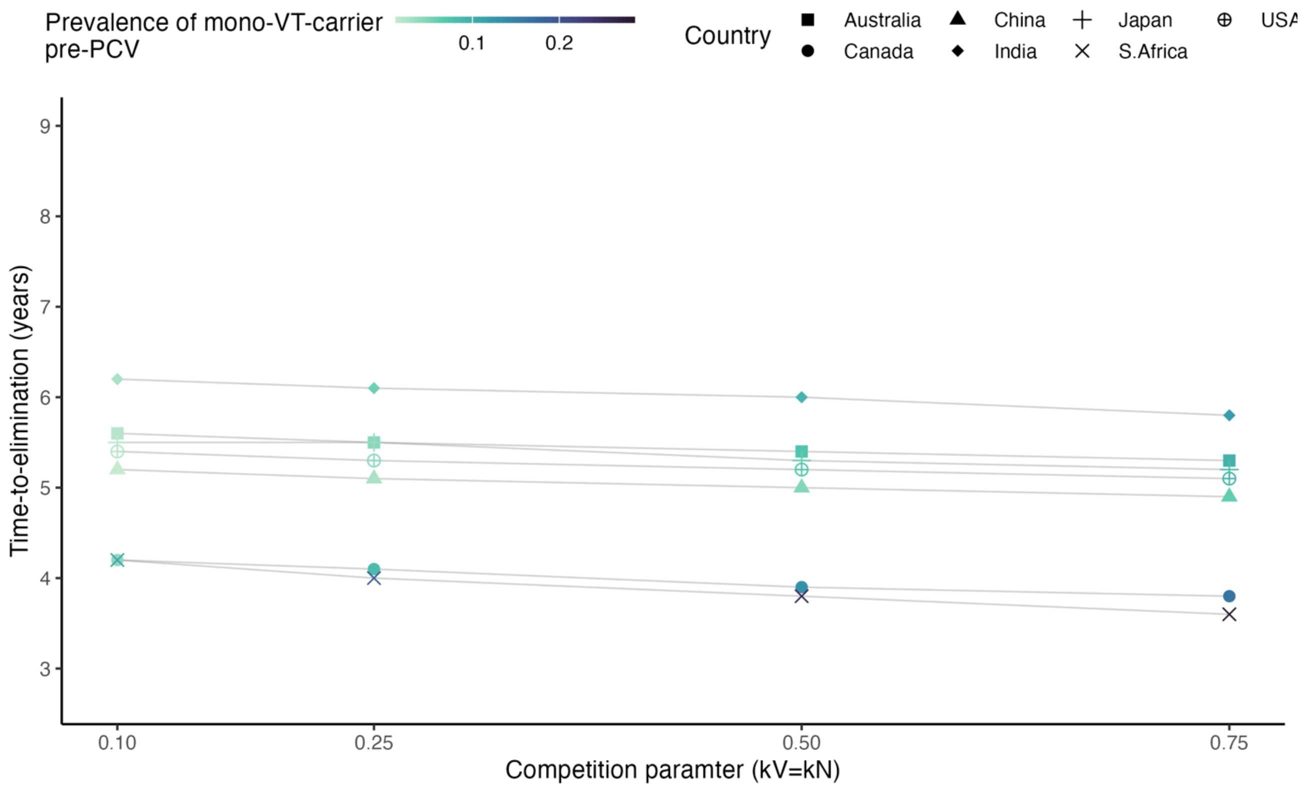


Keeping initial $F$ fixed at 0.65, we tested stronger ($k_V$=$k_N$=0.1, 0.25) and weaker ($k_V$=$k_N$=0.75) competitions. We found that time-to-elimination remained similar (time-to-elimination range: 4.3–6.3 years, 4.1–6.1 years, 3.6–5.8 years) to that in the main analysis (3.8–6 years).

The competition modelled here referred to the direct competition between VT and NVT in the nasopharynx, such that an existing colonization with VT (or NVT) would reduce colonization with NVT (or VT) by 25% ($k_V$=$k_N$=0.75), 50% ($k_V$=$k_N$=0.5), 75% ($k_V$=$k_N$=0.25), or 90% ($k_V$=$k_N$=0.1). As competition became stronger, co-carriers became less prevalent while mono-VT carriers and mono-NVT carriers became more prevalent, which led to a longer time for VT to be eliminated.

**Figure 9. Association of time-to-elimination and features of contact patterns in all age groups**

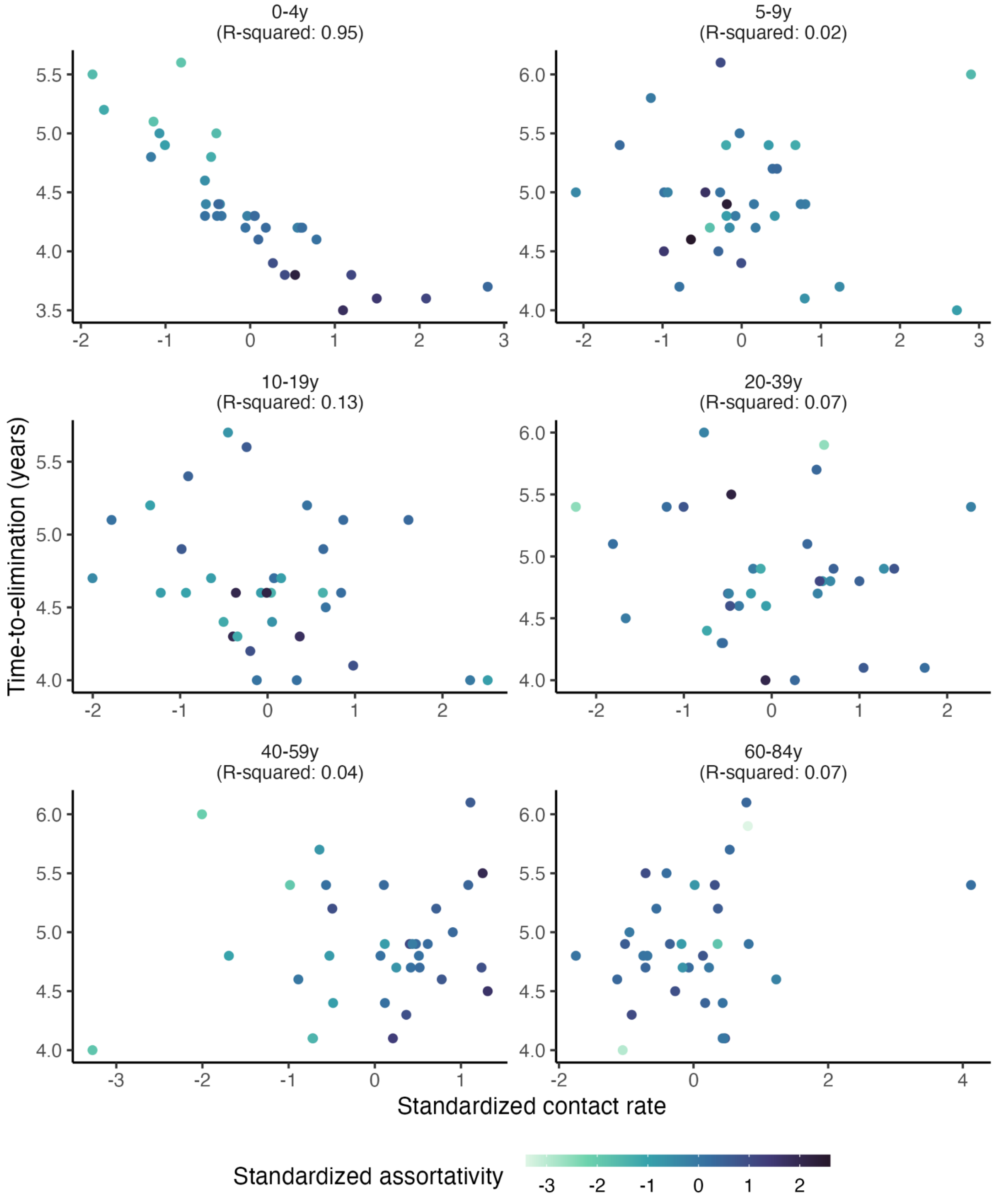


After simulating the transmission using the contact matrices from 34 countries in [1], we explored the relationship between contact features and time-to-elimination in each age group.

The time-to-elimination was measured for each age group. We then calculated the total number of daily contacts in an age group, divided by the age group width, such that this measure would not be inflated in the age groups of wider bands. For example, for age group 0–4y, the total number of daily contacts was divided by 5; for age group 20–39y, the total number of daily contacts was divided by 20. We defined assortativity as the average fraction of contacts from within the age group out of total contact for each age in the age group. We standardized both measures of contact features for easier comparison.

Figure 9 shows time-to-elimination's correlation with standardized contact rate (x-axis) and standardized assortativity (colour scale) for all age groups, revealing a strong trend in children under 5. The R-squared value in each panel indicates the variability in time-to-elimination explained by standardized contact rate and standardized assortativity in a generalized linear model (GLM) for each respective age group.

**Table 2. Out-of-sample prediction using contact rate and assortativity as predictors**

| Iteration | Contact matrices in the test set | MAE | MRAE |
|---|---|---|---|
| 1 | Czech, Finland, Bulgaria, Switzerland | 0.06 | 1.2% |
| 2 | Spain, Netherlands, Bulgaria, United Kingdom | 0.21 | 5.0% |
| 3 | Slovenia, Austria, Ireland, Lithuania | 0.07 | 1.7% |
| 4 | Australia, Norway, Sweden, Austria | 0.09 | 1.9% |
| 5 | Hungary, India, Bulgaria, Ireland | 0.19 | 3.7% |
| 6 | Netherlands, Bulgaria, Austria, India | 0.20 | 4.0% |
| 7 | Cyprus, South Africa, Hungary, United States | 0.06 | 1.5% |
| 8 | United States, Spain, Estonia, Portugal | 0.08 | 1.8% |
| 9 | Finland, Canada, Romania, Italy | 0.08 | 2.2% |
| 10 | France, Canada, Germany, Norway | 0.07 | 1.9% |

To test the predictive performance of the generalized linear model (GLM) using only standardized contact rate and standardized assortativity in children under 5 as predictors, we left 4 randomly selected contact matrices out as the test set and used the remaining 30 contact matrices as the training set. We repeated the procedure for 10 times and reported the mean absolute error (MAE) and mean relative absolute error (MRAE) for each iteration.

We found that a GLM containing only standardized contact rate and standardized assortativity in children under 5 predicted well the time-to-elimination in the test set, with MAE ranging from 0.06 to 0.21 years corresponding to 1.2–5% MRAE in the 10 iterations.

16. Hussain M, Melegaro A, Pebody RG, George R, Edmunds WJ, Talukdar R, et al. A longitudinal household study of Streptococcus pneumoniae nasopharyngeal carriage in a UK setting. Epidemiol

Infect. 2005;133: 891–898.
